# Rapid and Multiplexed Nucleic Acid Detection using Programmable Aptamer-Based RNA Switches

**DOI:** 10.1101/2023.06.02.23290873

**Authors:** Zhaoqing Yan, Anli A. Tang, Amit Eshed, Zackary M. Ticktin, Soma Chaudhary, Duo Ma, Griffin McCutcheon, Yudan Li, Kaiyue Wu, Sanchari Saha, Jonathan Alcantar-Fernandez, Jose L. Moreno-Camacho, Abraham Campos-Romero, James J. Collins, Peng Yin, Alexander A. Green

**Affiliations:** Department of Biomedical Engineering, Boston University, Boston, MA, USA; Molecular Biology, Cell Biology & Biochemistry Program, Graduate School of Arts and Sciences, Boston University, Boston, MA, USA; Biological Design Center, Boston University, Boston, MA 02215, USA; Biodesign Center for Molecular Design and Biomimetics at the Biodesign Institute, Arizona State University, Tempe, AZ, USA; School of Molecular Sciences, Arizona State University, Tempe, AZ, USA; Innovation and Research Department, Salud Digna, Culiacan, Sinaloa, Mexico; Department of Biological Engineering, Massachusetts Institute of Technology (MIT), Cambridge, MA, USA; Institute for Medical Engineering and Science, MIT, Cambridge, MA, USA; Wyss Institute for Biologically Inspired Engineering, Harvard University, Boston, MA, USA; Broad Institute of MIT and Harvard, Cambridge, MA, USA; Department of Systems Biology, Harvard Medical School, Boston, MA, USA; These authors contributed equally: Zhaoqing Yan, Anli A. Tang

## Abstract

Rapid, simple, and low-cost diagnostic technologies are crucial tools for combatting infectious disease. Here, we describe a class of aptamer-based RNA switches called aptaswitches that recognize specific target nucleic acid molecules and respond by initiating folding of a reporter aptamer. Aptaswitches can detect virtually any sequence and provide a fast and intense fluorescent readout, generating signals in as little as 5 minutes and enabling detection by eye with minimal equipment. We demonstrate that aptaswitches can be used to regulate folding of six different fluorescent aptamer/fluorogen pairs, providing a general means of controlling aptamer activity and an array of different reporter colors for multiplexing. By coupling isothermal amplification reactions with aptaswitches, we reach sensitivities down to 1 RNA copy/µL in one-pot reactions. Application of multiplexed one-pot reactions against RNA extracted from clinical saliva samples yields an overall accuracy of 96.67% for detection of SARS-CoV-2 in 30 minutes. Aptaswitches are thus versatile tools for nucleic acid detection that can be readily integrated into rapid diagnostic assays.

## INTRODUCTION

Fluorescent light-up aptamers have emerged as powerful tools for visualizing the dynamics of RNA expression in living cells^1–3^ and providing a bright, translation-free readout for *in vitro* reactions^4–6^. These systems consist of short sequences of RNA or DNA that bind to conditionally fluorescent dye molecules known as fluorogens to activate their fluorescence. Since the 2003 discovery of the MG RNA aptamer for binding malachite green^7^, diverse aptamer/fluorogen pairs have been developed with high binding specificity and varying spectral and photophysical properties^1^. In 2014, Jaffrey et al. reported the Broccoli aptamer that binds to the GFP-mimicking fluorogen DFHBI-1T and provides robust folding and green fluorescence in living cells^8^. This work was followed by the development of the Corn, Red Broccoli, and Orange Broccoli aptamers that bind to fluorogens to generate yellow, red/orange, and orange fluorescence, respectively^9, 10^. Unrau et al. have reported the Mango family of aptamers that bind to thiazole orange derivatives and offer brightness levels that significantly exceed enhanced GFP^11, 12^. These fluorogenic RNA aptamers have been used as tags for *in vivo* mRNA imaging and in biosensors for detecting proteins, small molecules, and nucleic acids^5, 13–17^.

Over the last decade, Ebola outbreaks in West Africa, the Zika epidemic, and the COVID-19 pandemic have highlighted the urgent need for rapid, sensitive, and low-cost diagnostics for containing the spread of viruses and ensuring that patients receive timely treatment^18, 19^. Nucleic-acid-based biomarkers associated with disease are essential for diagnostics because DNA and RNA can be amplified from trace amounts and provide highly specific detection. Conventional molecular assays, however, are implemented in centralized laboratories with expensive thermal cycling equipment and trained personnel, which increases costs, delays results, and reduces accessibility^20^. To reduce equipment needs, a variety of nucleic acid amplification methods that take place at a constant temperature have been developed. Unfortunately, these isothermal amplification strategies often suffer from false-positive results arising from non-specific amplification products. Thus, multiple approaches have been developed to verify the sequences of products following isothermal amplification while providing a visible readout. CRISPR-based diagnostic assays^20^ that employ cognate guide RNAs and collateral cleavage to verify the sequence of products and generate readout signals have been developed^21–25^. Similarly, paper-based cell-free transcription-translation systems have made use of sequence-specific riboregulators to confirm amplicon sequences down to the single-nucleotide level while providing a convenient color-based readout using protein reporters^19, 26–30^.

We hypothesized that sequence-specific RNA-based sensors employing light-up aptamer reporters could offer compelling advantages over previous sequence-verification methods for diagnostics. For instance, the rainbow of available fluorogenic aptamer colors could offer improved multiplexing capacity and target sequence versatility compared to CRISPR-based methods, which can have PAM site requirements and more limited multiplexing capacity. Moreover, the rapid folding of aptamers could enable much faster readout compared to diagnostics based on cell-free systems, whose readout speed is inherently limited by the rate of protein folding. Although a number of RNA-based sensors using aptamer reporters have been described, which range from split aptamer systems^3, 4, 31, 32^ to strand-displacement approaches^33, 34^, these systems have been limited in terms of dynamic range, activation speed, sequence versatility, and aptamer compatibility^3, 4, 31–36^.

Herein we report a new class of programmable aptamer-based RNA switches or aptaswitches that provide wide dynamic range, fast activation speeds, and multiplexing capacity. Aptaswitches exploit sequence-variable stems identified within a reporter aptamer structure to tightly repress its folding. Upon hybridization with a target nucleic acid sequence, aptaswitches exploit a toehold-mediated interaction mechanism to release this repression and promote fast aptamer folding, yielding wide dynamic range up to 260-fold and activation in as little as 5 minutes. Building on measurements of over a hundred aptaswitches, we implement a computational pipeline for rapid sensor design and demonstrate the generalizability of the aptaswitch mechanism by applying it to six different fluorogenic aptamer reporters having a variety of fluorescent colors. Coupling the aptaswitches with isothermal amplification provides clinically relevant sensitivity with fluorescence that can be seen by eye with minimal equipment. We further demonstrate multiplexed one-pot assays using the aptaswitches that detect extracted SARS-CoV-2 RNA from 60 clinical saliva samples in 30 minutes with an accuracy of 96.7%, highlighting the great utility of aptaswitches for rapid, low-cost diagnostic assays.

## RESULTS

### Broccoli aptaswitch design and validation

To implement a programmable aptamer-based switch, we first investigated the Broccoli RNA aptamer, which is known for its robust folding and bright fluorescence upon binding to the fluorogen DFHBI-1T^8^. Reasoning that an aptamer that could tolerate sequence changes would be easier to integrate into such a switch, we examined whether Broccoli could accommodate variations in its stem sequences without abolishing Broccoli/DFHBI-1T fluorescence. Two versions of Broccoli were evaluated: a ‘standard’ version in the same orientation originally reported by Filonov et al.^8^ and a ‘rotated’ version formed from a circular permutation of the aptamer that cleaved the aptamer at its apical loop and fused its original 3’ and 5’ ends within a loop (**Fig. 1a**). The base or ‘stabilizing’ stem of these versions of the aptamer, containing the 5’ and 3’ ends of the RNA, was then varied in sequence and tested for fluorescence with DFHBI-1T, while the core Broccoli sequence responsible for ligand binding was kept constant. Encouragingly, we found that both the standard and rotated forms of Broccoli retained strong fluorescence signals for all 16 different stem sequences tested (see **Supplementary Fig. S1a**).

**Fig. 1.**
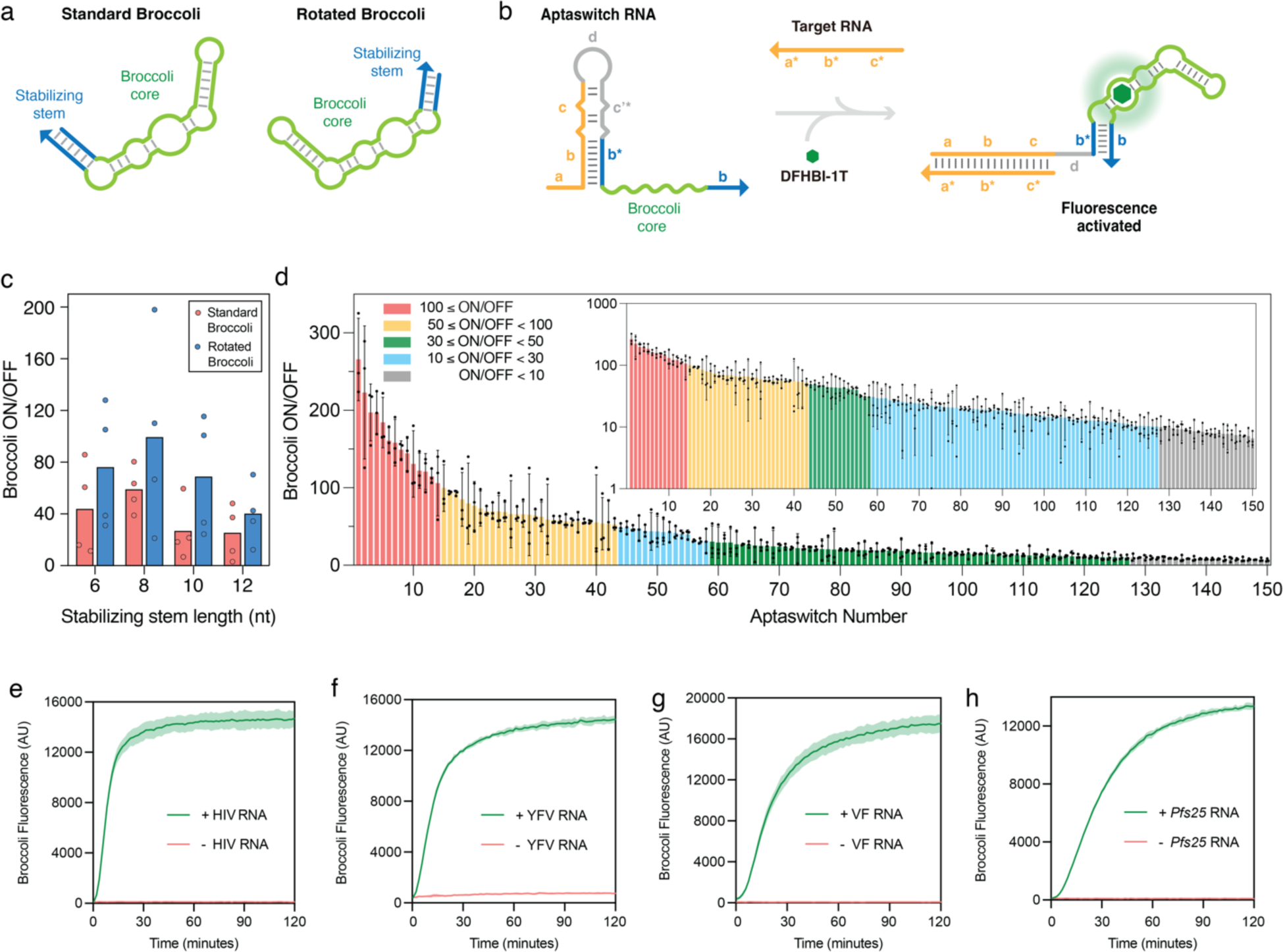
Mechanism of de novo-designed aptaswitches and *in vitro* characterization. **a**, Structures of the standard and rotated Broccoli aptamer. The standard Broccoli is the original Broccoli, and the rotated Broccoli is the circular-permutated Broccoli. **b**, Design schematics of Broccoli aptaswitches. Aptaswitches comprise: (i) a target-binding sequence that is complementary to a target nucleic acid or to the complement thereof (shown in yellow); and (ii) an aptamer core sequence (shown in green). The hairpin structure sequestering the stabilizing stem of the Broccoli represses the formation of the Broccoli/DFHBI-1T complex. The a* domain in the target RNA binds to a complementary, single-stranded toehold a domain in the aptaswitch RNA, initiating a branch migration that opens the hairpin stem. Newly released b* domain binds to the downstream b domain, enabling the formation of the Broccoli-DFHBI-1T complex. **c**, Evaluation of a library of 32 aptaswitches with standard and rotated versions of Broccoli having different stabilizing stem lengths. Data represent mean and individual ON/OFF values after 2 h of four aptaswitch designs with the same stabilizing stem domain length for each Broccoli version. **d**, ON/OFF fluorescence levels obtained 2 h after reaction for 150 first-generation Broccoli aptaswitches determined in the presence or absence of the cognate target RNA. Inset: ON/OFF GFP fluorescence measured for Broccoli aptaswitches on a logarithmic scale. Relative errors for the switch ON/OFF ratios were obtained by adding the relative errors of the switch ON and OFF state fluorescence measurements in quadrature. Relative errors for ON and OFF states are from the SD of *n*=3 technical replicates. **e-h**, Time-course measurements of fluorescence from top-performing Broccoli aptaswitches with and without the SARS-CoV-2 target RNA for the detection of HIV RNA (**e**), YFV RNA (**f**), VF RNA (**g**), Malaria-related *Pfs25* RNA (**h**). Shaded regions denote mean ± SD with *n*=3 technical replicates.

With this knowledge, we implemented the first-generation aptaswitch design shown in **Fig. 1b**. Each aptaswitch contains the full sequence of the reporter aptamer, including its core sequence and domains **b** and **b\*** encoding the stabilizing stem of the aptamer, where ‘*’ denotes a reverse complementary sequence. In the absence of the target RNA, however, the aptaswitch forces the aptamer into an inactive conformation by sequestering its **b\*** stem domain into a strong upstream hairpin structure. The 20-bp hairpin stem is substantially longer than the stabilizing stem, which discourages aptamer folding, and its 5’ positioning ensures that the hairpin can form prior to the aptamer during transcription, further reducing potential leakage. To ensure that the aptaswitches could respond rapidly to diverse target RNAs, we designed them to adopt a toehold-mediated strand displacement mechanism upon activation to provide fast reaction kinetics^37^ (**Fig. 1b**). Thus, each aptaswitch also contains a 15-nt 5’ single-stranded toehold domain **a** that is designed to initiate hybridization with a complementary target RNA. When the target RNA binds, it invades the hairpin stem, disrupting its structure and releasing the aptamer stem domain **b\***. To further encourage strand invasion, we incorporated a pair of single-nucleotide bulges into the upper stem of the hairpin in domains **c** and **c’\***, with **c** fully complementary to the target RNA, which increased the thermodynamic driving force of the forward reaction^38^ and decreased the likelihood of premature transcriptional termination. The stem domain **b\*** released from the sequestering hairpin can then hybridize with the exposed **b** domain and promote folding of the Broccoli reporter aptamer and generate fluorescence upon binding of DFHBI-1T. Importantly, this general aptaswitch design can be activated by diverse target RNA sequences, provided that the reporter aptamer has a stabilizing stem that can tolerate sequence variations.

We used the NUPACK nucleic acid sequence design package^39^ to generate initial libraries of *de*-*novo*-designed Broccoli aptaswitches in the standard and rotated configurations with a variety of target RNA sequences (**Fig. 1c**). The Broccoli aptaswitches routinely achieved greater than 40-fold ON/OFF ratios, which is defined as the ratio of the aptaswitch fluorescence with its cognate target divided by the fluorescence of the aptaswitch without it. Moreover, they showed that on average the rotated version of Broccoli provided better ON/OFF performance compared to the standard Broccoli. We thus proceeded to evaluate a library of over 150 first-generation aptaswitches using rotated Broccoli targeting a variety of RNAs taken from conserved regions of the genomes of 10 human pathogens: HIV, yellow fever virus (YFV), *Coccidioides* (causative agent of valley fever, VF), malaria-causing *Plasmodium falciparum* (antigen 25 mRNA, *Pfs25*), dengue virus (DENV), Chikungunya virus (CHIKV), St. Louis encephalitis virus (SLEV), West Nile virus (WNV), zika virus (ZIKV), and norovirus (NoV). Candidate Broccoli aptaswitches were designed for each potential binding site along the target RNA in 1-nt increments, leaving flanking regions on the 5’ and 3’ ends of the target available for primer binding. Sensors were designed to target either the sense or antisense orientation of the genome, which can both be readily generated following amplification, and the length of the stabilizing stem domain **b** was varied between 6 and 10 nt. The top six to eight constructs with the lowest overall defects for each target were tested experimentally. Sensor transcripts prepared by *in vitro* transcription were challenged with synthetic versions of the pathogen RNA targets. For determination of the ON/OFF ratio, background fluorescence from the DFHBI-1T fluorogen alone was not subtracted from either the ON- or OFF-state fluorescence.

**Figure 1d** shows the performance of the top 150 aptaswitches tested based on ON/OFF ratio. Overall, we found that 14 aptaswitches from this library provided a remarkable 100-fold or more increase in fluorescence upon detection of the cognate target and 43 provided a greater than 50-fold ON/OFF ratio. Time-course measurements of the optimal Broccoli aptaswitches from four pathogen target RNAs (**Fig. 1e-h**) demonstrated that the sensors can generate strong fluorescence signals within 15 minutes in 37°C reactions. The optimal Broccoli aptaswitch for the *P. falciparum* transcript *Pfs25* was also selected for target RNA titration experiments and found to activate with as little as 20 nM of target RNA (see **Supplementary Fig. 1c-d**).

### Second-generation aptaswitches enable multiplexed detection

To extend the utility of the aptaswitches, we sought to improve their performance using a second-generation design and enable multiplexed readout by integrating other fluorogenic aptamers. Based on the results from the library of first-generation Broccoli aptaswitches, we incorporated three design changes into the second-generation systems (**Fig. 2a**). First, we removed the bulges from the top of the hairpin and set its stem to be 6 bp longer than the stabilizing stem **b** domain of the reporter aptamer. This design change removed the possibility of leakage due to bulges in the stem, while reducing the average hairpin length to make it easier to transcribe and disrupt through strand invasion. Second, we allowed the toehold length of the aptaswitches to vary from 12 to 21 nt depending on the target RNA. Third, for reporter aptamers like Broccoli that have an apical stem loop that tolerates sequence changes, we allowed this stem-loop sequence to be optimized based on the larger context of the aptaswitch. This sequence was designed to promote folding of the aptamer, increasing ON-state signal, while minimizing potential for base pairing elsewhere within the aptaswitch, such as in the toehold regions. We then used computational RNA design to generate second-generation aptaswitches using six different aptamer/fluorogen pairs: Broccoli/DFHBI-1T, Red Broccoli/OBI, Orange Broccoli/DFHO, Corn/DFHO, Mango-III(A10U)/TO1-biotin, and Mango-IV/YO3-biotin. Corn and the Mango aptamers, which lack a variable apical stem loop, were designed with the updated toehold and hairpin alone. With the ongoing concern over mosquito-borne illnesses and the difficulties in identifying them^19, 40–42^, we targeted the aptaswitches to conserved genomic regions for these pathogens: CHIKV, DENV, *Pfs25* RNA from *P. falciparum*, WNV, YFV, and ZIKV.

**Fig. 2.**
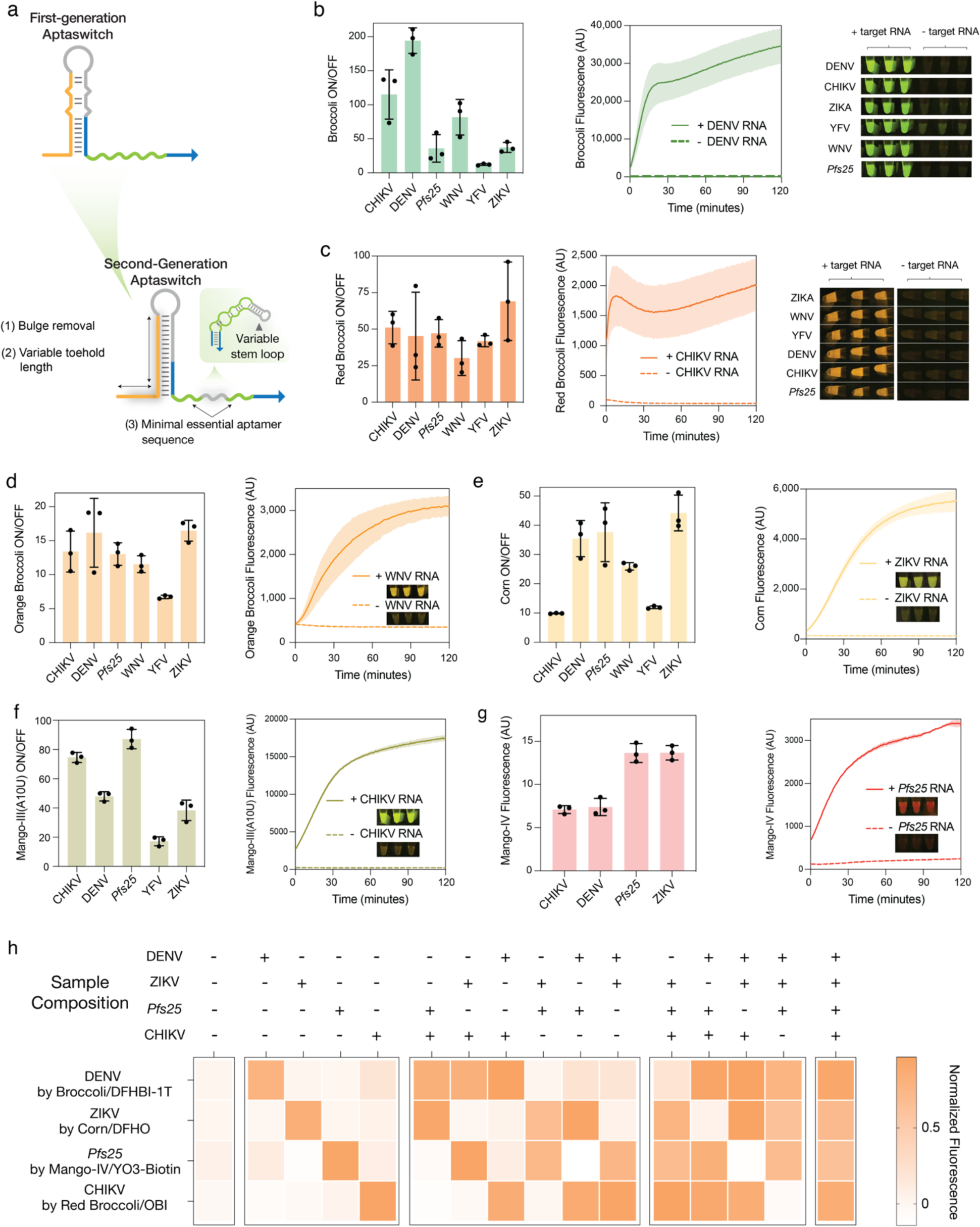
Second-generation aptaswitches enable multiplexed detection with orthogonal reporter aptamers. **a**, Schematic of the improved second-generation aptaswitch design compared to the first generation aptaswitches. **b-c,** ON/OFF fluorescence ratios, time-course measurements, and reaction photographs of Broccoli (b) and Red Broccoli (c) aptaswitches. **d-g,** ON/OFF fluorescence ratios and time-course measurements of Orange Broccoli (**d**), Corn (**e**), Mango-III(A10U) (**f**), and Mango-IV (**g**) aptaswitches. Inset: Photographs of aptaswitch reactions. **h**, Multiplexed detection of DENV RNA, ZIKV RNA, *Pfs25* RNA, and CHIKV RNA with Broccoli/DFHBI-1T, Corn/DFHO, Mango-IV/YO3-biotin, and Red Broccoli/OBI, aptamer/fluorogen pairs, respectively. The heat map represents the arithmetic mean of three replicates. For **b-g**, Bars represent the arithmetic mean ON/OFF ratio or fluorescence ± SD (*n*=3 technical replicates).

Using the second-generation design, we found that effective aptaswitches could be generated for all six reporter aptamers for the six pathogen target RNAs (**Fig. 2b-g**). The ON/OFF ratios of the aptaswitches varied depending on the target and aptamer, yet still provided significant differences in signal. Time-course measurements of representative aptaswitches from each reporter demonstrated that significant fluorescence is obtained from the sensors within 30 minutes, with some generating strong signals within 5 to 10 minutes (**Fig. 2b,c,f**). To observe the fluorescence of the aptaswitches, we illuminated the reactions using a blue-light transilluminator system equipped with an orange optical filter. The strong fluorescence produced by the aptaswitches with the cognate target was clearly visible using this setup (**Fig. 2b-g**). For potential in-home use, we found that inexpensive light and filter combinations costing about $23 could be used to make the aptaswitch reactions visible (**Supplementary Fig. 2a-c**). In addition, we found that the aptaswitch/fluorogen reactions are capable of being lyophilized and reactivated by adding water, suggesting potential for room-temperature storage and distribution (**Supplementary Fig. 2d**).

The ability to simultaneously detect multiple targets independently is highly desirable as it can be used to distinguish multiple illnesses that share similar symptoms at higher throughput and lower cost. Taking advantage of the distinct spectral properties of the aptamer/fluorogen pairs, we implemented several multiplexed reactions enabling independent detection of two to three different target RNAs for the mosquito-borne infections (**Supplementary Fig. 3a-i**). Leveraging the spectrally orthogonal Broccoli/DFHBI-1T, Corn/DFHO, Mango-IV/YO3-biotin, and Red Broccoli/OBI pairs, we implemented a four-channel reaction for simultaneous detection of targets from DENV, ZIKV, *Pfs25*, and CHIKV, respectively, which could successfully distinguish all 16 possible combinations of targets (**Fig. 2h**, see **Supplementary Fig. 3j** for photographs of the reactions).

### Integrating aptaswitches with isothermal amplification

We next focused on coupling the aptaswitches to isothermal amplification reactions to ensure that they could reach clinically relevant detection limits for pathogen nucleic acids. We first tested the aptaswitches in combination with nucleic acid sequence-based amplification (NASBA)^43^ reactions that operate at 41°C for detection of DENV (**Supplementary Fig. 4**). NASBA followed by aptaswitch readout enabled detection down to sample concentrations of 2.41 RNA copies/µL (**Supplementary Fig. 4d**) and allowed us to correctly identify three DENV-positive and three DENV-negative serum samples (**Supplementary Fig. 4e**). However, the two-hour reaction time needed for NASBA reactions prompted us to explore more rapid isothermal amplification methods.

Reverse transcription loop-mediated isothermal amplification (RT-LAMP) is a fast and cost-effective method to amplify specific RNA sequences within minutes^44, 45^. RT-LAMP has an operating temperature of ∼61°C to 71°C and uses a set of four to six primers to generate DNA products containing exposed, single-stranded loop domains^46, 47^. We sought to use the aptaswitches in combination with RT-LAMP to develop an assay to detect SARS-CoV-2 ^48^, which has accounted for more than 670 million infections and more than 6 million deaths worldwide^49^. We thus developed aptaswitches that targeted the exposed loop domains of the RT-LAMP products to activate aptamer fluorescence (**Fig. 3a**). The use of aptaswitches reduces the potential for false positive results by ensuring that the correct sequence is generated from RT-LAMP, a common failure mode for assays that employ isothermal amplification alone^20^.

**Fig. 3.**
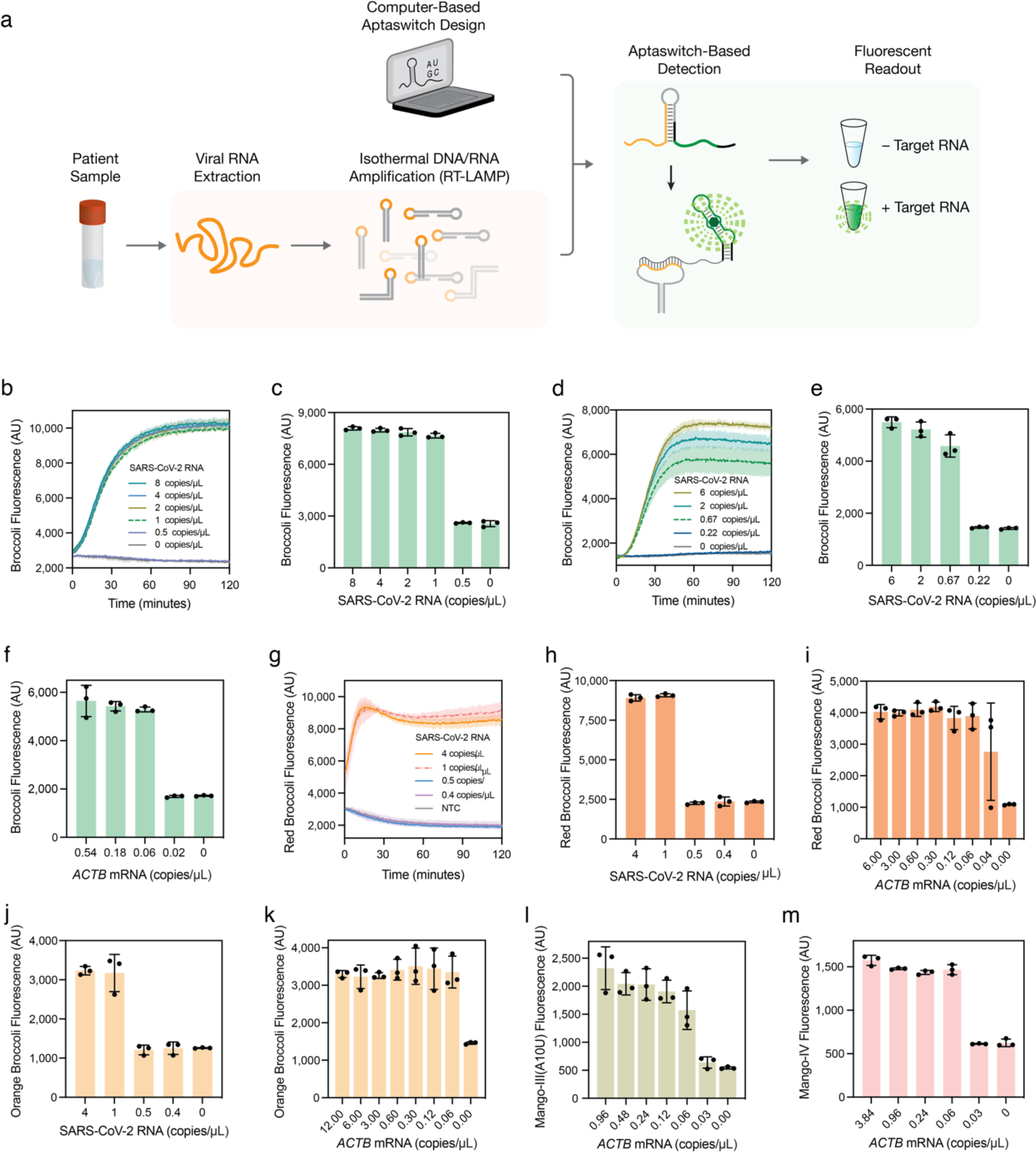
Integration of Isothermal amplification with aptaswitches to enable detection of attomolar concentrations of viral RNA. **a**, Schematic of the general procedure for detection of viral RNA. RNA is extracted from patient samples and amplified using RT-LAMP. Amplified nucleic acids are then detected using specific aptaswitches. The aptaswitch binds to exposed loop domains in the RT-LAMP DNA amplicons to produce the output fluorescence signal. A strong fluorescent signal is then used to indicate that viral RNA is present in the patient sample. **b**-**f**, Representative fluorescence plots of Broccoli aptaswitches for detection of RT-LAMP amplification for N gene (**b**) and S gene (**d**) of RNA from cultured SARS-CoV-2. **c**, **e**, and **f** show bar graphs of analytical sensitivity determination using series diluted cultured SARS-CoV-2 RNA (**c** and **e**) or synthetic human control *ACTB* mRNA (**f**). Bars in **c**, **e**, and **f** represent the mean fluorescence intensity ± SD of the aptaswitches at 30 min, which was preceded by a 30-minute RT-LAMP reaction. **g**-**i**, Limit of detection for Red Broccoli aptaswitches for the detection of SARS-CoV-2 N gene (**h**) and *ACTB* mRNA (**i**). **g** shows the Red Broccoli aptaswitch response following RT-LAMP with different template concentrations as a function of time. (*n*=3 technical replicates; bars represent the arithmetic mean ± SD) **j**-**k**, LoD for Orange Broccoli aptaswitches for the detection of SARS-CoV-2 N gene (**j**) and human control ACTB mRNA (**k**). (*n*=3 technical replicates; bars represent the arithmetic mean ± SD) **l**-**m**, Fluorescence intensity measured for different human control ACTB mRNA concentrations after 30 mins of Mango-III(A10U) (**i**) and Mango-IV aptaswitch (**m**). (*n*=3 technical replicates; bars represent the arithmetic mean ± SD)

We first designed second-generation aptaswitches targeting multiple regions of the SARS-CoV-2 genome and human β-actin (*ACTB*) mRNA, which serves as a sample processing control, using six different reporter aptamers: Broccoli, Red Broccoli, Corn, Mango-III(A10U), Mango-IV, and Orange Broccoli. Initial aptaswitch screens were performed using DNA stem-loop sequences that mimicked the expected RT-LAMP amplicons. We observed impressive signal increases greater than 100-fold with rapid system activation against these targets for the Broccoli aptaswitches (**Supplementary Fig. 5a-e** with DFHBI-1T fluorogen and **Supplementary Fig. 5f-j** with BI fluorogen) and the Red Broccoli/OBI aptaswitches (**Supplementary Fig. 6a-c**) against SARS-CoV-2 and *ACTB*. Aptaswitches using Corn/DFHO, Mango-III(A10U)/TO1-biotin, Mango-IV/YO3-biotin, and Orange Broccoli/DFHO combinations were also effective at detecting both types of targets albeit at lower optimal ON/OFF ratios between 20-to 50-fold (**Supplementary Fig. 6d-n**).

The top-performing aptaswitches from these screens were coupled to RT-LAMP in two-pot assays using different combinations of primers and aptaswitches to gauge their sensitivity. In these two-pot reactions, RT-LAMP is first performed on the RNA sample for 30 minutes at 61°C, and the resulting RT-LAMP product is then diluted into a second reaction at 37°C containing the aptaswitches designed to target the amplicon loop domains. Since RT-LAMP also produces amplicons with two loop regions that have unrelated sequences, we also implemented a scheme to target the two independent loop regions in the same reaction (**Supplementary Fig. 7a**). This approach effectively doubles the concentration of targets available for detection and thus increases signal output and/or decreases reaction time (**Supplementary Fig. 7b-d).** A pair of Broccoli aptaswitches targeting two RT-LAMP loops from amplification of the N gene and S gene provided a limit of detection of 1 copy/µL and 0.67 copies/µL, respectively, in the amplification reaction and activated within 30 minutes (**Fig. 3b-e**). Using RT-LAMP primers amplifying the human control *ACTB* mRNA and a pair of Broccoli aptaswitches, we achieved an impressive limit of detection of 0.06 copies/µL in the amplification reaction (**Fig. 3f**). Red Broccoli aptaswitches provided red/orange fluorescence, could activate within 15 minutes, and demonstrated detection limits of 1 copy/µL and 0.06 copies/µL for the SARS-CoV-2 RNA (N gene) and *ACTB* mRNA, respectively, using the same sets of RT-LAMP primers (**Fig. 3g-i**). Orange Broccoli, Mango-III(A10U), and Mango-IV were also successfully integrated into the two-pot reaction scheme and provided the same limits of detection as the Broccoli and Red Broccoli sensors (**Fig. 3j-m**). Experiments performed with these aptamer outputs rapidly provided a visible color that could be seen by the eye using a blue light source and an orange optical filter (**Supplementary Fig. 8**).

To increase the throughput and ease of implementation of the tests, we also developed master mix formulations for reactions that can be stably stored at -20°C and rapidly added to 384-well plates at the time of use (**Supplementary Fig. 9**). The parallelized two-pot 384-well assay employs a 30-minute RT-LAMP reaction, followed by dilution into a 25-minute aptaswitch detection reaction. The assay was tested against contrived samples containing extracted RNA from SARS-CoV-2, other human coronaviruses, and influenza at concentrations typical of clinical saliva samples. The high-throughput aptaswitch assay successfully identified the SARS-CoV-2 samples and provided excellent specificity in under one hour of total reaction time (**Supplementary Fig. 9c**).

### Developing an aptaswitch-based all-in-one molecular diagnostic assay

One-pot diagnostic assays where all reaction steps occur in the same reaction vessel and do not require additional reagents to be added after the start of the reaction are highly desirable. Such assays reduce processing time, labor, time to result, and the likelihood of cross-contamination, while providing increased throughput. Accordingly, we developed a streamlined one-pot RT-LAMP/aptaswitch assay for detection of SARS-CoV-2 RNA where the reaction is heated 30 minutes for amplification, followed by a return to room temperature for aptaswitch readout (**Fig. 4a**). To implement the assay, we found that it was essential to choreograph binding of the LAMP primers and aptaswitches to the amplicons at different reaction stages to avoid assay inhibition. During the amplification phase, we used a raised 67°C reaction temperature (**Supplementary Fig. 10a**) and a higher primer/aptaswitch ratio (**Supplementary Fig. 10b**) to promote primer binding over aptaswitch binding. Once amplification is complete and most of the primers are consumed, the return to room temperature facilitates aptaswitch binding, aptamer folding, and fluorogen docking to generate a strong fluorescence signal (**Fig. 4a**, **Supplementary Fig. 10c**). We found that standard aptaswitch buffer components and the fluorogen inhibited RT-LAMP reactions (**Supplementary Fig. 10d**). Thus, we kept the fluorogen concentrations low and did not add additional chemical components to the reactions and determined that both RT-LAMP and aptaswitch readout worked effectively in these conditions (**Supplementary Fig. 10d**).

**Fig. 4.**
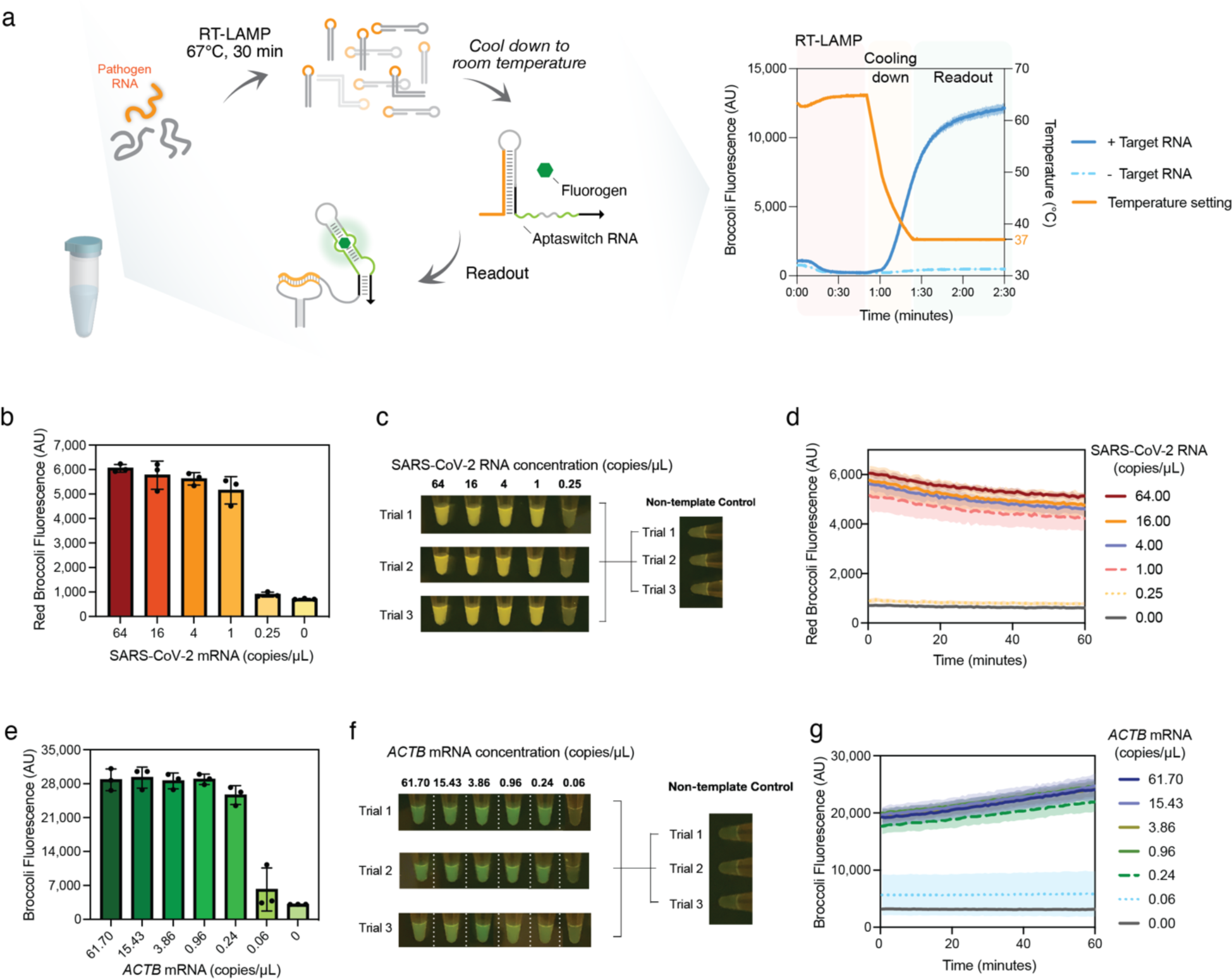
Adapting aptaswitches for one-pot RT-LAMP reactions. **a**, Schematic of target RNA detection by one-pot isothermal amplification-coupled aptaswitch. A 30-minute amplification step at 67°C is used for the RT-LAMP reaction and followed by cooling to room temperature. Over this second stage, the reduced temperature enables the aptaswitches to bind to the RT-LAMP amplicons and the cognate fluorogen. A strong and rapid increase in fluorescence is observed in the reaction signaling the presence of target viral RNA. **b**-**d**, Analytical sensitivity of one-pot Red Broccoli aptaswitch for the detection of SARS-CoV-2 N gene using different cultured SARS-CoV-2 RNA template concentrations (**b**). **c** shows photograph of fluorescence from triplicate Red Broccoli aptaswitch reactions following the 30-minute amplification step at 67°C. Time-course measurements obtained from a plate reader of aptaswitch with different target RNA concentrations at 25°C (**d**). Bars (**b**) and shaded regions (**d**) denote the arithmetic mean ± SD with *n*=3 technical replicates. **e**-**g**, Fluorescence intensity from Broccoli aptaswitch reactions immediately following the 30-minute amplification step at 67°C for the detection of human control ACTB mRNA using different synthetic RNA template concentrations (**e**). **f** shows photograph of fluorescence from Broccoli aptaswitch reactions at room temperature. Time-course measurements record fluorescence intensity from Broccoli aptaswitch reactions from a plate reader at 25°C (**g**). Reactions were measured in triplicate. Bars (**e**) and shaded regions (**g**) denote the arithmetic mean ± SD with *n*=3 technical replicates.

In the optimized one-pot assays, RNA extracted from a patient sample is added to a reaction combining RT-LAMP components, along with the aptaswitches and fluorogens for target detection. The reaction vessel is first heated to 67°C for 30 minutes to amplify the genetic material from the pathogen followed by cooling to room temperature, where the aptaswitches can bind to the amplicon loops. We first tested the one-pot reactions using the optimal Red Broccoli aptaswitches targeting SARS-CoV-2. These experiments revealed that the Red Broccoli aptaswitches could provide visible fluorescence output for sample concentrations down to 1 RNA copy/µL in the reaction (**Fig. 4b-c**). In addition, strong fluorescence was observed from the aptawitches by the time the system reached room temperature (**Fig. 4d**). A pair of Broccoli aptaswitches also successfully detected the SARS-CoV-2 S gene in a one-pot assay using a temperature-controlled plate reader to provide fluorescence readout in real time (**Supplementary Fig. 10e**). In addition, we deployed a pair of Broccoli aptaswitches targeting two loops of the RT-LAMP amplicon of the *ACTB* mRNA for detection of the control transcript in a one-pot reaction (**Fig. 4e-g**). Once again, Broccoli fluorescence was detectable by the time the reaction reached room temperature (**Fig. 4f-g**) and yielded a detection limit of 0.24 copies/µL of *ACTB* mRNA.

We next took advantage of the multiplexing capabilities of the aptaswitches to apply to the one-pot RT-LAMP/aptaswitch reactions to detect SARS-CoV-2 and the *ACTB* control mRNA simultaneously (**Fig. 5a**). This multiplexed reaction employed 12 different primers from the two sets of RT-LAMP primers, Broccoli and Red Broccoli aptaswitches, and the DFHBI-1T and OBI fluorogens needed for each reporter aptamer. While developing the assay, we found that the Red Broccoli aptaswitches, which favor binding to OBI, also bound to DFHBI-1T in the reaction conditions needed for the one-pot reactions. This effect led to a strong green fluorescence signal along with the desired red/orange fluorescence upon activation of Red Broccoli. Consequently, we chose to use a Red Broccoli aptaswitch for detection of SARS-CoV-2 amplicons and a pair of Broccoli aptaswitches for detection of the *ACTB* amplicon. This combination of sensors still allowed for accurate results to be obtained. Negative clinical samples would generate green fluorescence from the presence of the *ACTB* mRNA in the sample via Broccoli/DFHB-1T, confirming a successful reaction. For samples positive for SARS-CoV-2, a red/orange signal from Red Broccoli/OBI would indicate both the presence of the SARS-CoV-2 amplicon and a successful reaction, obviating the need for readout from the *ACTB* aptaswitch.

**Fig. 5.**
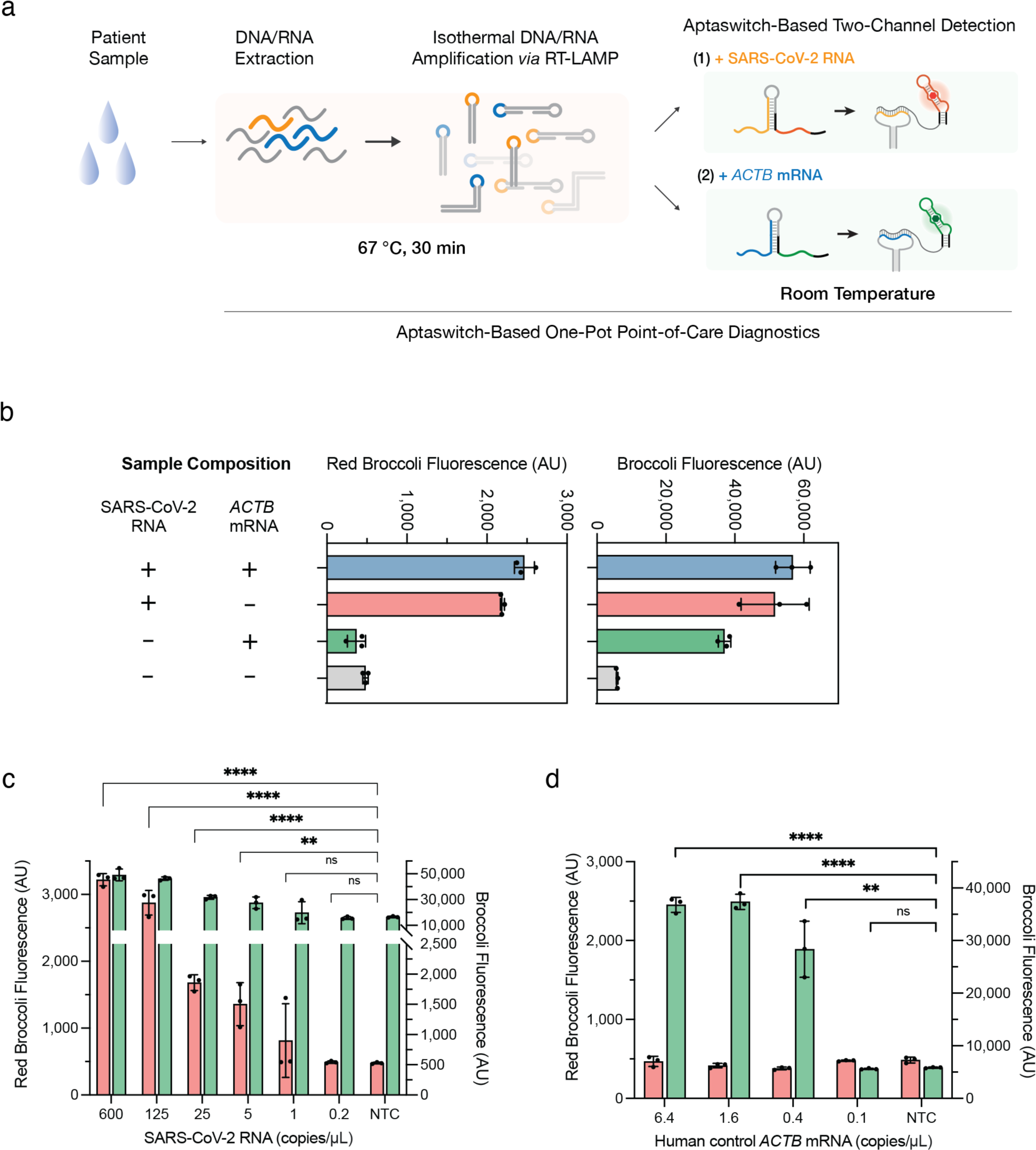
Multiplexed one-pot RT-LAMP/aptaswitch assay for detection of viral RNA. **a**, Schematic of the multiplexable one-pot RT-LAMP/aptaswitch assay. RT-LAMP is initially performed at 67°C for 30 minutes for simultaneous amplification of RNA targets, followed by a cooling period. Reduction of temperature enables binding of the aptaswitches and is evidenced by a rapid increase in the fluorescence of both Broccoli and Red Broccoli aptaswitches, signifying the presence of both the SARS-CoV-2 N gene and the *ACTB* mRNA. **b**, One-pot simultaneous detection of SARS-CoV-2 RNA and human control *ACTB* mRNA with Red Broccoli and Broccoli aptaswitches, respectively. (*n*=3 technical replicates; bars represent the arithmetic mean ± SD) **c**, Limit of detection assay of for SARS-CoV-2 N gene using 4000 copies/μL *ACTB* mRNA. Values are mean ± SD with *n*=3 technical replicates. Red bar represents fluorescence intensity of Red Broccoli aptaswitches for detecting SARS-CoV-2 RNA. Green bar represents fluorescence intensity of Broccoli aptaswitches for detecting ACTB mRNA. (Two-tailed Student t test; ns, p > 0.05; **, p < 0.01; ***, p < 0.001; ****, p < 0.0001.) **d**, Limit of detection assay of one-pot multiplexed aptaswitches for *ACTB* mRNA without SARS-CoV-2 RNA. (*n*=3 technical replicates, two-tailed Student t test; ns, p > 0.05; **, p < 0.01; ***, p < 0.001; ****, p < 0.0001. Values are the arithmetic mean ± SD. Red bar: fluorescence intensity of Red Broccoli aptaswitches for detecting SARS-CoV-2 RNA; green bar: fluorescence intensity of Broccoli aptaswitches for detecting *ACTB* mRNA).

We initially tested the multiplexed one-pot assay using contrived samples with different combinations of the SARS-CoV-2 and *ACTB* synthetic target RNAs and found that it generated the expected two-channel fluorescence signals (**Fig. 5b**). Once the system is activated, the signal remains stable at room temperature over six days (**Supplementary Fig. 11a-b**). To assess the sensitivity of the multiplexed one-pot reactions, we prepared synthetic samples with SARS-CoV-2 RNA concentrations varying from 600 copies/µL to 0.2 copies/µL while the *ACTB* mRNA concentration was kept constant at 4000 copies/µL in the reaction (**Fig. 5c**). A significant Red Broccoli fluorescence was observed in the presence of the SARS-CoV-2 N gene down to concentrations of 5 copies/µL, while the assay control green fluorescence remained high for all *ACTB*-containing samples. The assay was then tested by supplying different concentrations of *ACTB* mRNA in the absence of SARS-CoV-2 (**Fig. 5d**). These experiments demonstrated that the one-pot multiplexed RT-LAMP/aptaswitch assay could provide significant fluorescence output down to 0.4 RNA copies/µL of the *ACTB* mRNA in the reaction.

### Validation of multiplex one-pot reactions with clinical saliva samples

We next sought to validate the assay using a panel of 31 SARS-CoV-2 positive and 29 negative clinical saliva samples. RNA from the samples was first extracted using a PureLink RNA extraction kit and SARS-CoV-2 RNA concentrations were quantified via reverse transcription-quantitative polymerase chain reaction (RT-qPCR). The extracted RNA was supplied to the multiplexed one-pot aptaswitches/RT-LAMP reactions and incubated at 67°C for 30 minutes followed by rapid cooling for readout in 384-well plates using a plate reader. We used the 95^th^ percentile value for non-template controls of 26 valid (*ACTB* mRNA present) and 31 invalid samples (*ACTB* mRNA not present) for threshold value determination of SARS-CoV-2 RNA and human control gene assay channels (**Supplementary Fig. 12a**). These thresholds from Red Broccoli aptaswitches for determining the presence of SARS-CoV-2 RNA and Broccoli aptaswitches for determining the presence of human control *ACTB* mRNA were then applied to the analysis of data from double-blinded testing of patient saliva samples. Analysis of the samples following reaction cool down enabled immediate identification of positive and negative samples in a plate reader. **Figure 6a-b** and **Supplementary Fig. 12b** show the fluorescence of the Red Broccoli and Broccoli aptaswitches at the beginning of the measurement. In general, the SARS-CoV-2 positive samples display a much higher Red Broccoli fluorescence than the negative samples as expected, while the Broccoli fluorescence remains highly active in all the samples. Applying the fluorescence thresholding criteria, the assay identified 29 out of 31 positive samples, which had Ct values ranging from 7 to 30 correctly, while 29 out of 29 negative samples were correctly determined to be free of SARS-CoV-2 RNA. Accordingly, the sensitivity of this assay 93.55% and the specificity is 100%, resulting in an overall accuracy of 96.67%.

**Fig. 6.**
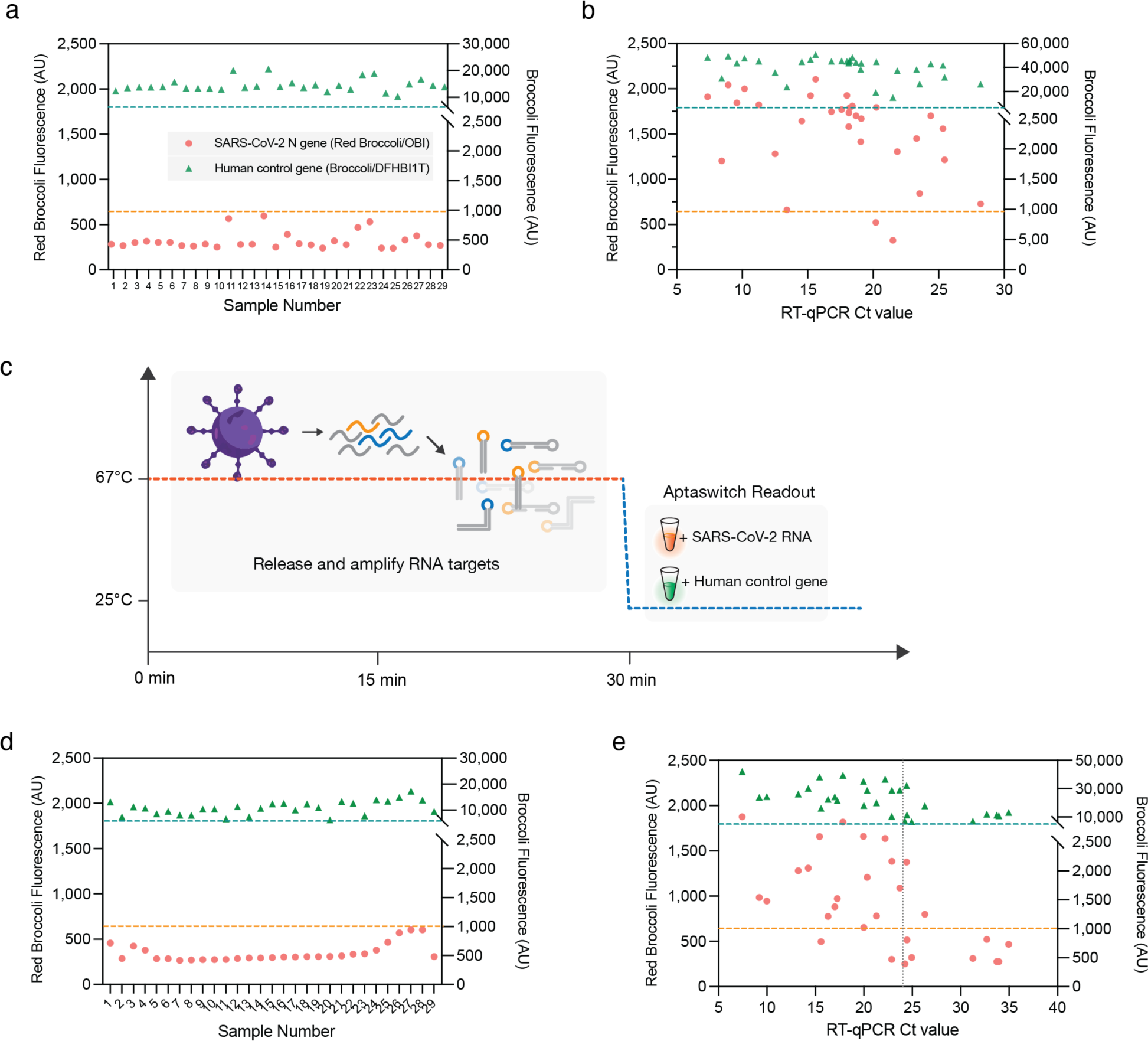
Validation of aptaswitches on clinical saliva samples. **a-b**, Fluorescence signals obtained from extracted RNA from clinical saliva samples for (**a**) 29 negative and (**b**) 31 positive clinical saliva samples were measured. The dashed lines represent diagnostic thresholds that were determined using 95^th^ percentile values for non-template controls of valid and invalid samples (orange dashed line: Red Broccoli aptaswitch threshold value for determining a SARS-CoV-2 positive sample; teal dashed line: Broccoli aptaswitch threshold value for determining a valid sample based on the presence of the human *ACTB* mRNA). **c**, Schematic of one-pot extraction-free detection of SARS-CoV-2 RNA in clinical saliva samples. The release of RNA from patient saliva samples at 67 °C along with RT-LAMP amplification in the same reaction. **d-e**, Fluorescence signals obtained from extraction-free multiplexed one-pot RT-LAMP/aptaswitch reactions for (**d**) 29 negative and (**e**) 29 positive clinical saliva samples. The dashed lines represent diagnostic thresholds that were determined using 95^th^ percentile values for non-template controls of SARS-CoV-2 positive and negative samples (orange dashed line: Red Broccoli aptaswitch threshold value for determining a SARS-CoV-2 positive sample; teal dashed line: Broccoli aptaswitch threshold value for determining a valid sample).

To investigate the potential of the multiplexed one-pot assay for rapid high-throughput testing and in-home use, we also evaluated the assay without the RNA extraction step. In this streamlined workflow, the saliva sample is directly added to the RT-LAMP/aptaswitch mixture and the 67°C reaction temperature is used to simultaneously release viral RNA from the SARS-CoV-2 capsid and amplify its genetic material (**Fig. 6c**). A total of 58 clinical saliva samples were analyzed using this approach (**Fig. 6d-e** and **Supplementary Fig. 12c**). For the negative patient samples, 29 out of 29 were identified correctly (**Fig. 6d**). However, we found that for positive patient samples with Ct > 24 only 2/10 were identified correctly (**Fig. 6e**). This reduction in sensitivity may have been caused by components from saliva interfering with the RT-LAMP efficiency. Despite this limitation, we found that the one-pot direct-from-saliva assay successfully identified 17 out of 19 positive patient samples with higher viral loads (Ct ≤ 24). For these higher viral load saliva samples, the direct assay yielded an accuracy of 95.8% with 89.5% sensitivity and 100% specificity. While the performance of the direct-from-saliva assay is substantially below that of RT-qPCR tests, the ease of implementation and fast 30-minute results from these one-pot tests is advantageous for frequent at-home testing. Moreover, the use of saliva samples, which are collected non-invasively and do not require swabs, makes the direct assay very attractive compared to rapid antigen tests, which typically demonstrate poor sensitivity when used with saliva^50–52^.

## DISCUSSION

We have developed a class of computer-designed RNA aptaswitches enabling detection of target nucleic acids without any sequence constraints using a variety of different fluorogenic aptamers for readout. Demonstrating the broad adaptability and programmability of this approach, we assembled a set of over 700 aptaswitches capable of targeting diverse RNAs, including pathogens and human transcripts. Taking advantage of the orthogonal aptamer reporters, we implemented a four-channel single-reaction multiplexing detection scheme capable of detecting DENV, ZIKA, Malaria, and CHIKV targets simultaneously. This capability can be used to reduce assay costs and processing time. Combining aptaswitches for detection with isothermal reactions for amplification, we achieved clinically relevant levels of sensitivity for viral detection in two-pot reactions. Use of aptaswitches reduces the potential for false positive results by ensuring that the correct sequence is generated from isothermal amplification, a common failure mode for assays that employ isothermal amplification alone. We found using RT-LAMP that the sensitivity of the assay is equivalent to RT-qPCR tests conventionally used for SARS-CoV-2 detection in clinical samples. Clear fluorescence signals from the assays can be visualized using simple, readily available equipment that can be obtained for about $23.

Finally, we also demonstrated one-pot multiplexed SARS-CoV-2 detection with a human control gene through RT-LAMP/aptaswitch reactions using Red Broccoli/OBI and Broccoli/DFHBI-1T in combination to obtain robust test results within 30 minutes. This assay was able to reliably detect as few as 5 copies/μL of SARS-CoV-2 with red/orange fluorescence from Red Broccoli, while simultaneously providing readout of the assay control *ACTB* mRNA using the Broccoli fluorescence channel. The assay could be extended to higher-order multiplexing with other common respiratory viruses, such as Influenza A or respiratory syncytial virus (RSV) using additional orthogonal aptaswitch/fluorogen pairs offering distinct fluorescence signatures.

Key to the high performance and sequence versatility of the aptaswitches is the exploitation of a variable stem region within the reporter aptamers to regulate their folding. Harnessing this feature, which occurred at one or two sites for all fluorogenic aptamers we tested, enabled us to repress or activate aptamer folding using virtually any sequence. The repression mechanism we use based on disruption of the stabilizing stem, in turn, enables the aptaswitches to be activated quickly with high yields since only a single intramolecular hybridization reaction is necessary to form the aptamer. This direct activation mechanism simplifies the design and the aptamer folding pathway compared to other approaches requiring an additional strand-displacement step for activation^34^. The resulting optimized repression and activation mechanisms lead to fast turn-on speed and wide dynamic range, with ON/OFF ratios reaching 260-fold for the best aptaswitches. Furthermore, use of a sequestration mechanism that hides key components of the aptamer, namely the stabilizing stem, within the sensors contributes to the programmability of the aptaswitches. Sequestration means that fewer nucleotides of the aptamer are exposed and only the toehold region of the target-binding site is available for binding, which reduces the likelihood of off-target hybridization and sensor misfolding. In contrast, split aptamer systems, where the reporter aptamer is cleaved in two for repression, employ fully exposed target-binding regions^3, 4, 31, 32^. These exposed regions can lead to sensor misfolding based on the target sequence, hampering recognition of the target molecule and limiting sensor programmability.

Taken together, our all-in-one multiplexed aptaswitch assay is sensitive, low-cost, rapid, and easy-to-use, and it reduces the need for centralized and expensive instrumentation and diagnostic professionals. We anticipate our aptamer-based molecular diagnostic system will enable individuals to on-site test more frequently with a shortened turn-around time^53, 54^. Our success in using *in silico* design tools to reliably produce high-performance, specific aptaswitches reduces the number of design cycles required for optimization by improving the components tested with each passing cycle. Thus, multiplexed aptaswitches can also be quickly engineered for profiling custom or newly emerging pathogens. We expect that future improvements will enable the use of aptaswitches to resolve sequences down to the single-nucleotide level, opening the door to rapid genotyping and variant identification. Furthermore, use of alternative aptamers and substrates should enable the aptaswitches to be used in colorimetric assays that require no additional optical equipment. The success of our aptaswitch with multiple fluorogenic aptamers suggests that they can be broadly applied to aptamers in general and in other contexts. For instance, such systems could be used in living cells for tag-free imaging of endogenous transcripts or for regulating the folding of non-fluorescent aptamer outputs.

## MATERIALS AND METHODS

### Transcriptional template preparation and RNA synthesis

All DNA oligonucleotides were purchased from Integrated DNA Technologies. DNA fragments were assembled and amplified via PCR using the Phusion High-Fidelity PCR Master Mix with HF Buffer (NEB, M0531L). PCR product constituting the transcriptional template for the aptaswitches was purified using a MinElute PCR Purification Kit (Qiagen, 28006).

Aptaswitch RNAs were *in vitro* transcribed at 37 °C for 2 hours using AmpliScribe™ T7-Flash™ Transcription Kits (Lucigen, ASF3507) with 200 nM of the DNA template. Synthetic target RNAs were synthesized from PCR-amplified templates and were purified using Monarch® RNA Cleanup Kit (NEB, T2040L). All transcription products were prepared using this method unless otherwise noted. For quantification of target RNA concentrations, DNase I (Lucigen, ASF3507) was used to remove the DNA template to terminate transcription.

### Experimental screening of aptaswitches

Six to eight promising aptaswitches identified during *in silico* selection were tested for each pathogen target. A BioTek Synergy Neo2 multimode microplate reader was used for all plate reader measurements. 1.0 μM of column-purified aptaswitches (for Mango-III(A10U) and Mango-IV aptaswitches) or 0.7 μL direct transcription product of aptaswitch RNA (for Broccoli, Corn, Red Broccoli and Orange Broccoli aptaswitches) and 2.0 μM of purified target RNA were added to the 384-well plate along with 4 μM of DFHBI-1T for Broccoli (Lucerna, 410), 4 μM of BI for Broccoli (Lucerna, 600), 2 μM of DFHO for Corn and Orange Broccoli (Lucerna, 500), 3 μM of OBI for Red Broccoli (Lucerna, 610), 2 μM of TO1-3PEG-Biotin for Mango-III(A10U) (abm, G955), or 2 μM of YO3-3PEG-Biotin for Mango-IV (abm, G957). DFHBI-1T, BI, OBI, TO1-3PEG-Biotin or YO1-3PEG-Biotin buffer consisted of 40 mM HEPES pH 7.4, 100 mM KCl, and 5 mM MgCl_2_. DFHO buffer consisted of 40 mM HEPES pH 7.4, 100 mM KCl, and 1 mM MgCl_2_. Before each measurement, samples were shaken linearly for 30 seconds to ensure proper mixing. The plate reader was preheated, and the measurements were taken at 37°C unless otherwise indicated. To visually observe the fluorescence of the sensors, we illuminated the reactions in a microplate using a Safe Imager 2.0 Blue Light Transilluminator (ThermoFisher, G6600).

### Multiplexed detection using orthogonal aptaswitches

Two-channel aptaswitch reactions were prepared with either a 2 μM concentration of the purified Corn aptaswitch or 1.4 μL of the direct transcription product of the Corn aptaswitch along with 1 μM DFHO fluorogen, and 1 μM of purified Broccoli aptaswitch or 0.7 μL of the direct transcription product of the Broccoli aptaswitch along with 4 μM of the DFHBI-1T fluorogen. The 10X fluorogenic dye mix is consisted of 10 μM DFHO, 40 μM DFHBI-1T, 40 mM HEPES pH 7.4, 100 mM KCl, and 5 mM MgCl_2_.

Three-channel aptaswitch reactions were prepared with 2 μM purified Corn aptaswitch along with 1 μM DFHO, 1 μM purified Broccoli aptaswitch along with 4 μM DFHBI-1T, and 1 μM purified Mango-IV aptaswitch along with 2 μM YO3-biotin. The 10X fluorogenic dye mix consisted of 10 μM DFHO, 40 μM DFHBI-1T, 20 μM YO3-biotin, 40 mM HEPES pH 7.4, 100 mM KCl, and 5 mM MgCl_2_.

Four-channel aptaswitch reactions were prepared with 2 μM purified Corn aptaswitch along with 1 μM DFHO, 1 μM purified Broccoli aptaswitch along with 4 μM DFHBI-1T, 1 μM purified Mango-IV aptaswitch along with 2 μM YO3-biotin, and 1 μM purified Red Broccoli aptaswitch along with 4 μM DFHBI-1T. The 10X fluorogenic dye mix consisted of 10 μM DFHO, 40 μM DFHBI-1T, 20 μM YO3-biotin, 40 μM OBI, 40 mM HEPES pH 7.4, 100 mM KCl, and 5 mM MgCl_2_.

Multiplexed reactions were incubated at 37°C for 30 min and transferred to a 384-well plate for fluorescence measurements in a Synergy Neo2 multimode microplate reader. The photographs of fluorescence were taken using a Safe Imager 2.0 Blue Light Transilluminator (Thermo Fisher, G6600).

### Analysis of fluorescence data from four-channel multiplexed aptaswitch reactions

Since the fluorescence of the reporter aptamers had some spectral overlap, we implemented a MATLAB script to deconvolve plate reader data and extract the fluorescence generated by each aptamer/fluorogen pair in the four-channel multiplexed reactions. A calibration plate was prepared that contained the aptaswitch reaction buffer with the four fluorogens in one well, representing a blank sample for background fluorescence subtraction, and four other wells containing the fluorogens in buffer with one of the aptamers. The five calibration wells were then measured in a Synergy Neo2 multimode microplate reader using four excitation/emission pairs: 472 nm/507 nm, 505 nm/545 nm, 595 nm/620 nm, 541 nm/590 nm. Following subtraction of the background signal, the characteristic profile of the four excitation/emission signals associated with each of the four reporter aptamers was determined, generating a 4×4 non-singular matrix linking the fluorescence reads to each aptamer. We inverted this matrix to generate a conversion matrix that could take fluorescence read data and convert it into the signal generated by each of the four reporter aptamers. Multiplexed reactions were first measured using identical conditions in the plate reader. Background-subtracted fluorescence from the reactions was then converted into aptamer signals using the conversion matrix (**Fig. 4h**). We also tested the accuracy of the conversion matrix by applying it to all possible combinations of two or more of the aptamers. Comparison of the known aptamer combinations to those generated by the conversion matrix from fluorescence reads yielded an R-squared value of 0.972, confirming the accuracy of the conversion matrix.

### NASBA reactions with aptaswitch readout

For target RNA amplification with NASBA, 2.01 μL 3X reaction buffer (Life Sciences, NEC-1-24;), 0.99 μL 6X nucleotide mix (Life Sciences, NEC-1-24), 0.03 μL Protector RNase Inhibitor (Roche), 0.12 μL of each NASBA primer (12.5 mM, IDT), 0.15 μL nuclease-free water (Thermo Fisher, 10977015), and 1.2 μL target RNA were mixed at 4°C and heated to 65°C for 2 min, followed by a 10-minute incubation at 41°C. 1.5 μL Enzyme Mix (Life Sciences, NEC-1-24) was then added to the reaction for a final volume of 6 μL. After mixing, the reaction was incubated at 41°C for 2 hours. For a 35 μL two-pot reaction, 5 μL of the NASBA amplified RNA product was combined with 0.7 μL of RNA aptaswitch and 10X DFHBI-1T dye mix. This reaction was incubated and measured at 37 °C for an additional 2 hr. The 10X DFHBI-1T dye mix consists of 40 μM DFHBI-1T (Lucerna, 410), 40 mM HEPES (Gibco™, 15630080), pH 7.4, 100 mM KCl (Invitrogen™, AM9640G), and 5 mM MgCl_2_ (Invitrogen™, AM9530G).

### Viral RNA extraction for clinical dengue virus samples

De-identified serum samples positive and negative for dengue virus were obtained at Salud Digna (Culiacan, Mexico). Viral RNA was extracted by using a QIAamp Viral RNA Mini Kit (Qiagen, 52904) according to the manufacturer’s instructions. RNA was eluted with 60 µL Buffer AVE (Qiagen, 1020953) and stored at -80°C before use.

### Two-pot RT-LAMP/aptaswitch reactions

RT-LAMP reactions were conducted with WarmStart LAMP 2X Master Mix (NEB, E1700L), 3 μL of 10X primer mix (IDT; 16 μM of FIP/BIP, 4 μM FLoop/BLoop, and 2 μM of F3/B3), and 12 μL of sample. The final volume of the amplification reaction was 30 μL. Amplification was performed at 61 °C for 30 min in an Applied Biosystems™ ProFlex™ PCR System (Thermo Fisher, 4484073). Heat-Inactivated 2019 Novel Coronavirus (ATCC, VR-1986HK^TM^) was used as a template to perform the aptaswitch screen and establish LOD ranges. The volume of template to be used and the final concentration of template in the reaction was calculated on the basis of the initial concentration provided by the vendor of 4.2×10^5^ copies/μL. 10 μL of the resulting RT-LAMP product was then added to the 384-well plate containing 0.7 μL of directly transcribed aptaswitch RNA (for Broccoli, Corn, Red Broccoli, or Orange Broccoli aptaswitches) along with 4 μM of DFHBI-1T (Lucerna, 410), 2 μM of DFHO (Lucerna, 500), 4 μM of OBI (Lucerna, 610), or 4 μM of DFHO (Lucerna, 500), respectively. For Mango aptaswitches, 1 μM of purified aptaswitch RNA was used with 2 μM of TO1-3PEG-Biotin (abm, G955), or 2 μM of YO3-3PEG-Biotin (abm, G957).

### One-pot one-channel RT-LAMP/aptaswitch reactions

The one-pot one-channel reaction combines RT-LAMP components [WarmStart LAMP 2X Master Mix (NEB, E1700L) and 3 µL 10× primer mix (IDT; 24 μM of FIP/BIP, 6 μM FLoop/BLoop and 3 μM of F3/B3)] along with the aptaswitches and corresponding fluorogen. These reactions used primers at 1.5x the standard concentrations for RT-LAMP (i.e. 2.4 µM for FIP and BIP, 0.3 µM for F3 and B3, and 0.6 µM for LF and LB primers). For Broccoli aptaswitch readout, the reactions contained 2 µM DFHBI-1T and 0.2 µL of the direct transcription products of each of the Broccoli aptaswitches targeting the forward and backward loops of the SARS-CoV-2 N gene RT-LAMP amplicon. For Red Broccoli readout, the reactions contained 2 µM OBI and 0.4 µL of the direct transcription product of the Red Broccoli aptaswitch targeting the backward loop of the ACTB mRNA RT-LAMP amplicon. No additional ions were added. Following addition of the SARS-CoV-2 sample, the reaction was incubated at 67°C for 30 min in a multiwell plate in a temperature-controlled plate reader with the fluorescence read out in real time or incubated in a thermal cycler or incubator. Following the 67°C incubation, the temperature of reaction was cooled to room temperature for aptaswitch binding and fluorescence readout.

### One-pot two-channel RT-LAMP/aptaswitch reactions

The one-pot two-channel RT-LAMP/aptaswitch assay was conducted with WarmStart LAMP 2X Master Mix (NEB, E1700L), 3 μL of 10X primer mix, 1.5 μL of 20X fluorogen mix, 0.8 μL of aptaswitch mix, and 9.7 μL of sample. The final volume of the amplification reaction was 30 μL. The 10X RT-LAMP primer mix includes two sets of primers: set 1 for the SARS-CoV-2 N gene at 1.5x the standard RT-LAMP concentration (IDT; 24 μM of FIP/BIP, 6 μM FLoop/BLoop and 3 μM of F3/B3) and set 2 for the ACTB mRNA control at 0.8x the standard RT-LAMP concentration (IDT; 12.8 μM of FIP/BIP, 3.2 μM FLoop/BLoop and 1.6 μM of F3/B3). The 20X fluorogen mix is composed of 40 μM OBI (Lucerna, 610) and 40 μM DFHBI-1T (Lucerna, 410). No additional ions were added. The aptaswitch mix consists of 2 µL of Broccoli aptaswitch for the ACTB mRNA amplicon F-loop, 2 µL of Broccoli aptaswitch for the ACTB mRNA amplicon B-loop, and 4 µL of Red Broccoli aptaswitch for the SARS-CoV-2 amplicon B-loop. The reaction was incubated at 67°C for 30 minutes, followed by a cooling period to 25 °C in an Applied Biosystems™ ProFlex™ PCR System (Thermo Fisher, 4484073), a temperature-controlled plate reader, or an incubator. The reactions were transferred to a 384-well plate for fluorescence measurements in a Synergy Neo2 multimode microplate reader. Broccoli/DFHBI-1T fluorescence was measured with 472 nm excitation and 507 nm emission wavelengths. Red Broccoli/OBI fluorescence was measured with 541 nm excitation and 590 nm emission wavelengths.

### Viral RNA extraction for clinical SARS-CoV-2 human saliva samples

Clinical saliva samples were obtained from the Biodesign Institute Clinical Testing Laboratory at Arizona State University. These specimens were collected for SARS-CoV-2 diagnostic purposes. The viral RNA was extracted by using a PureLink^TM^ Viral RNA/DNA Mini Kit (Thermo Fisher, 12280050) according to the manufacturer’s instructions. RNA was eluted with 50 µL of H_2_O and stored at -80°C before use.

### RT-qPCR measurements of clinical samples

RT-qPCR parallel detection was prepared using a Luna® SARS-CoV-2 RT-qPCR Multiplex Assay Kit Detection (NEB, E3019). The detection was performed following the protocol of the kit in 7900HT Fast Real-Time PCR System with a 384-well block module (Thermo Fisher, 4329001). Time to threshold was calculated using single-threshold analysis mode.

## Data Availability

The main data supporting the results in this study are available within the paper and the Supplementary Figures. The datasets generated during and/or analysed during the current study are available from the corresponding author on reasonable request.

## Acknowledgments

We thank Biodesign Institute Clinical Testing Laboratory at Arizona State University for providing COVID-19-relevant patient samples. We also thank Zhenwei Zhou (Boston University) for helpful discussions on the use of statistical methods for threshold value determination of SARS-CoV-2 diagnostic. This work was also supported by an NIH Director’s New Innovator Award (1DP2GM126892), an NIH U01 award (1U01AI148319-01), an NSF RAPID award (2029532), the Gates Foundation (OPP1160667), Arizona Biomedical Research Centre funds (ADHS16-162400, CTR051763), an Alfred P. Sloan Fellowship (FG-2017-9108), an NIH R21 award (1R21AI136571-01A1), and Canadian Food Inspection Agency funds (39903-200137) to A.A.G, along with funding from Salud Digna Research Council (SDI-20166).

## Author contributions

Z.Y. and A.A.T. performed most of the wet lab experiments, design of the sensors and experiments, selection of viral target sequences, and the analysis of data. Z.Y. conceived and developed second-generation, RT-LAMP-coupled, one-pot, and multiplexed aptaswitch assays and portable home setup for visualizing the signal. A.A.T. designed and performed NASBA-coupled aptaswitch experiments for detecting dengue virus from patients’ serum. J.A.-F., J.L.M.-C., A.C.-R. acquired, analyzed, and identified the dengue virus clinical serum samples. A.A.G. conceived and conceptualized the use of aptamer, generated the automatic codes of design of sensors and primers used for some of the assays. Z.Y. and A.A.G. interpreted data and wrote the manuscript. Z.Y. and A.A.T. developed the rapid sensor screening procedure. Z.Y., A.A.T, A.E., Z.M.T., D.M., Y.L., and K.W. performed the experiments. Z.M.T., S.C., and S.S. contributed to patient sample collection. G.M. optimized the design code and generated aptaswitch designs computationally. A.A.G., P.Y., and J.J.C. supervised the research.

