## Supplementary Figures for "Rapid and Multiplexed Nucleic Acid Detection using Programmable Aptamer-Based RNA Switches"

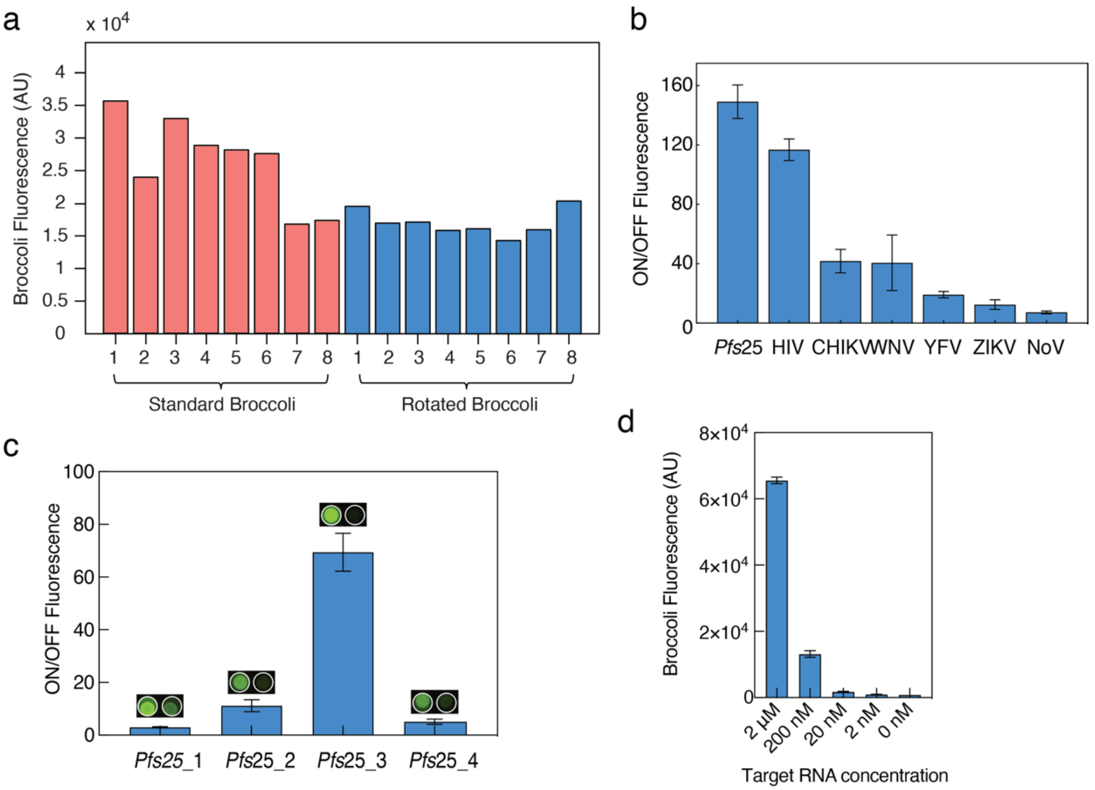


**Supplementary Fig. 1 | Parameter optimization of first-generation Broccoli aptaswitch designs.**

**a**, Fluorescence of the standard and rotated versions of the Broccoli aptamer with various stem sequences. Standard Broccoli displayed wider variations in fluorescence as a function of sequence. Rotated Broccoli fluorescence was less variable and generally lower than standard Broccoli, potentially due to its stem being one-nucleotide shorter.

**b**, The best-performing Broccoli aptaswitches targeting RNAs for each pathogen species. Data represent the ON/OFF fluorescence ratio from the sensors in the presence or absence of the cognate target RNA. Bars represent the arithmetic mean ± SD from *n*=3 technical replicates.

**c**, Screening of the Broccoli aptaswitches targeting the *pfs*25 gene of malaria. Photographs of fluorescence from the aptaswitch in the presence or absence of the cognate target RNA are shown above the bars. Samples were excited under a blue light transilluminator using a blue-light filter to remove excess excitation light.

**d**, Sensitivity of *Pfs*25_3 aptaswitch without RNA amplification. Bars represent the arithmetic mean ± SD from *n*=3 technical replicates.

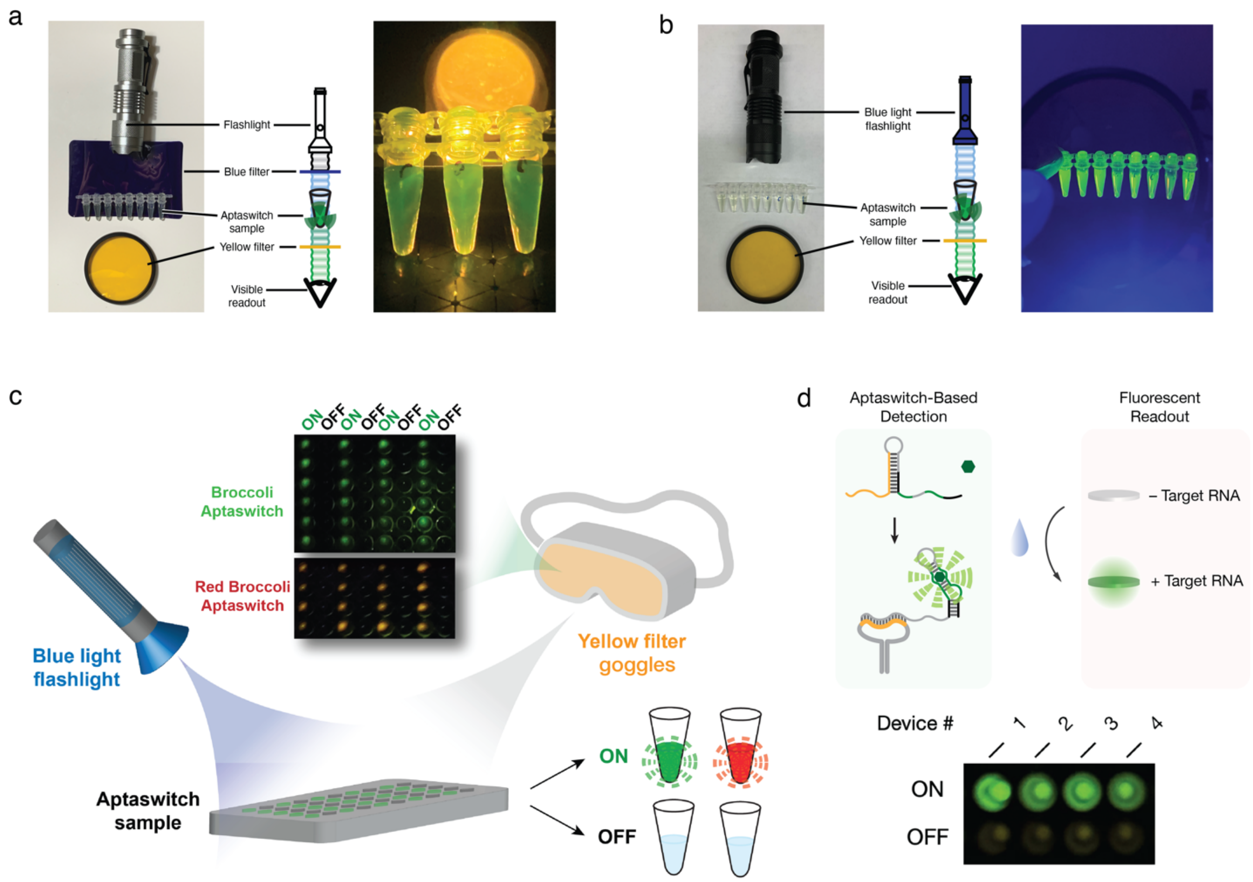


**Supplementary Fig. 2 I Simple, inexpensive home setups for readout of aptaswitch reactions by eye or on paper substrates.**

**a**, The first setup includes an LED Flashlight (7W 300 LM Mini LED, Wayllshine: $6.99), a blue filter (Pieces Universal Gels Lighting Filter Kit, Selens: $0.60/filter), and a yellow filter (Tiffen 58mm 12 Filter (Yellow): $14.82) that can be configured for detection of fluorescence from the Broccoli aptaswitches. Photograph of active Broccoli aptamers displaying green fluorescence using the setup.

**b**, The second setup includes a blue light flashlight (Blue LED 3 Mode Flashlight, Wayllshine: $8.99), while retaining the same yellow filter ($14.82) that can be configured for detection of fluorescence from the Broccoli aptaswitches. Photograph of active Broccoli aptamers displaying green fluorescence using the setup.

**c**, The third setup is suitable for high-throughput readout of aptaswitch reactions and includes a blue light flashlight and widely available yellow filter goggles for filtering out UV light (Calabria

1003 Large Fit-Over UV Protection in Yellow, $12.95). Photographs of active Broccoli and Red Broccoli aptamers displaying green and red fluorescence, respectively, using the setup. Negative samples lacking both targets displayed low fluorescence signals as expected.

**d**, Schematic of freeze-dried paper-based aptaswitch reactions for portable diagnostics. Photograph of active Broccoli aptamers displaying green fluorescence emission after a solution containing the target RNA is used to rehydrate the paper-based aptaswitch reactions.

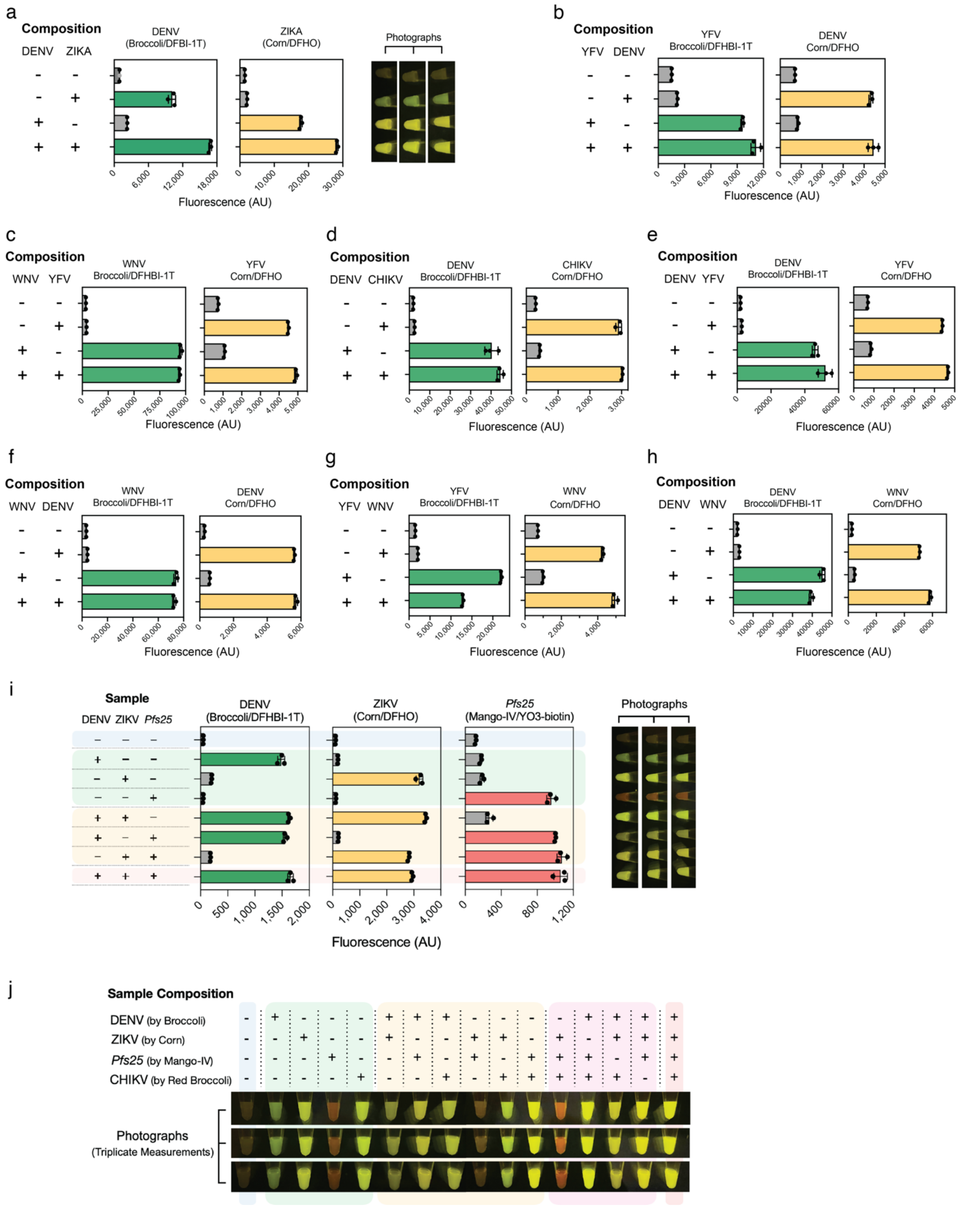


**Supplementary Fig. 3 | Multiplexed detection of up to four mosquito-borne pathogen target RNAs with spectrally distinct aptamer/fluorogen pairs.**

**a-h**, Two-channel simultaneous detection of different pairs of Mosquito-borne infections using Broccoli/DFHBI-1T and Corn/DFHO. Bars represent the arithmetic mean ± SD from *n*=3 technical replicates.

**i**, Three-channel multiplexed detection of DENV RNA, ZIKV RNA, and *Pfs25* RNA of *P. falciparum* with Broccoli aptaswitch/DFHBI-1T, Corn aptaswitch/DFHO, and Mango-IV aptaswitch/YO3-biotin, respectively. Bars represent the arithmetic mean ± SD from *n*=3 technical replicates.

**j**, Photograph of fluorescence from the four-channel multiplexed aptaswitch reactions measured in triplicate for DENV RNA, ZIKV RNA, *Pfs25* RNA of *P. falciparum*, and CHIKV RNA with Broccoli, Corn, Mango-IV, and Red Broccoli aptamer reporters, respectively.

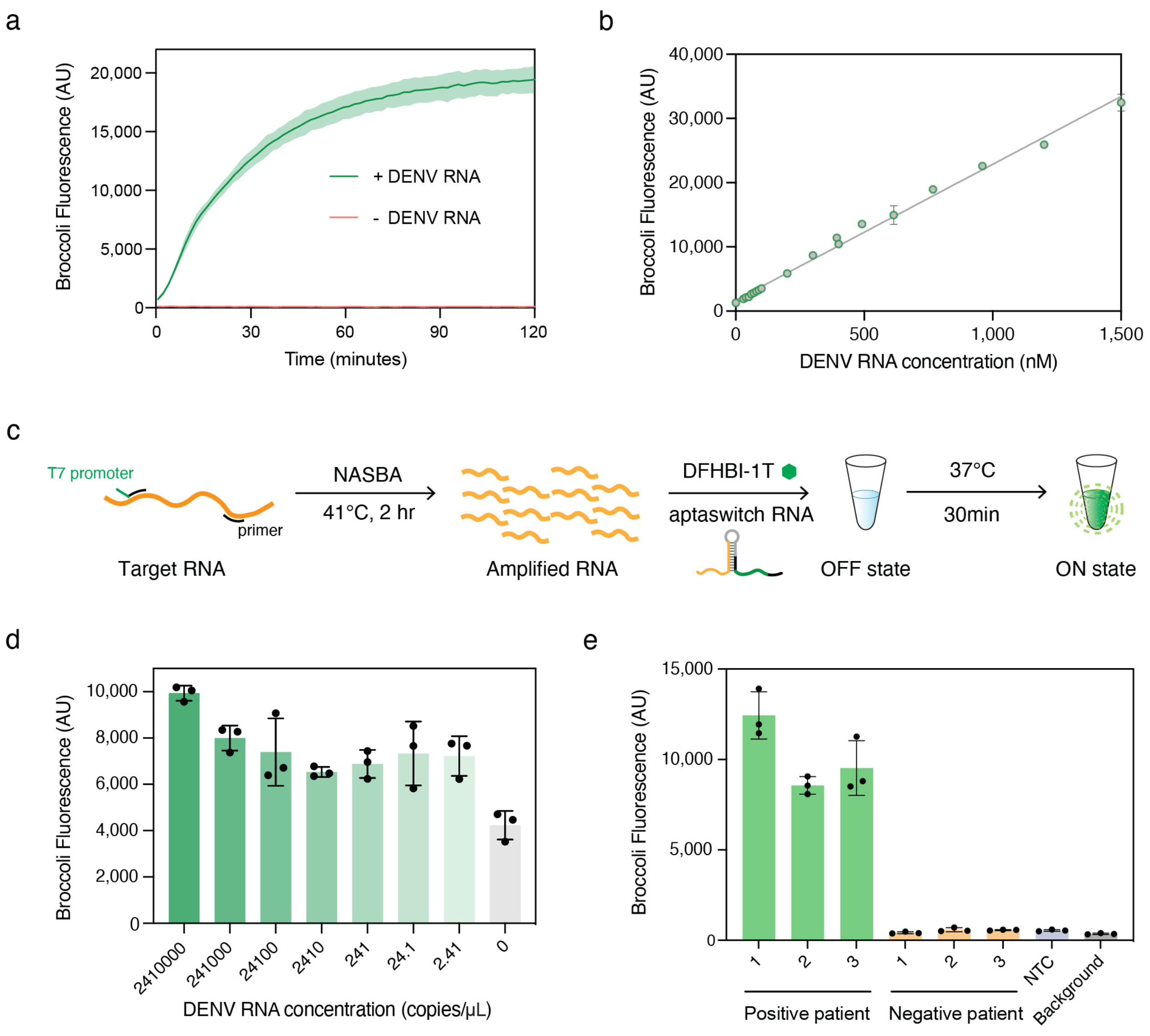


**Supplementary Fig. 4 l Coupling Broccoli aptaswitches with NASBA reactions for dengue virus detection.**

**a**, Optimal Broccoli aptaswitch for detecting dengue. Curves and shaded regions represent the arithmetic mean ± SD from $n=3$ biological replicates for aptaswitch ON and OFF states.

**b**, Calibration curve for aptaswitch-based detection of synthetic dengue virus target RNA. Aptaswitches displayed a linear response to target RNA concentration. Data points and error bars are the arithmetic mean ± SD from $n$=3 technical replicates.

**c**, Workflow for detecting pathogen RNA using a two-pot NASBA/Broccoli aptaswitch assay. NASBA is an enzymatic amplification process that is run in isothermal conditions and utilizes three enzymes: reverse transcriptase, RNase H, and T7 RNA polymerase. NASBA reactions were incubated at 41°C for 2 hours. The resulting amplicons were then added to solutions containing the Broccoli aptaswitch and DFHBI-1T. The fluorescence intensity of the aptaswitches upon binding to the NASBA products after 30 minutes is then used to indicate if dengue viral RNA is present in the patient sample.

**d**, Detection limit measurements for synthetic DENV target RNA subject to NASBA amplification and detection using Broccoli-based aptaswitch. A detection limit of 2.41 copies/$\mu$L in the amplification reaction was obtained. Bars represent the arithmetic mean ± SD from $n$=3 biological replicates.

**e**, DENV detection using viral RNA extracted from patient serum. Aptaswitch reactions were run on NASBA products generated after two hours of amplification. Aptaswitch test results agreed with those obtained using RT-qPCR. Bars represent the arithmetic mean ± SD from $n$=3 technical replicates.

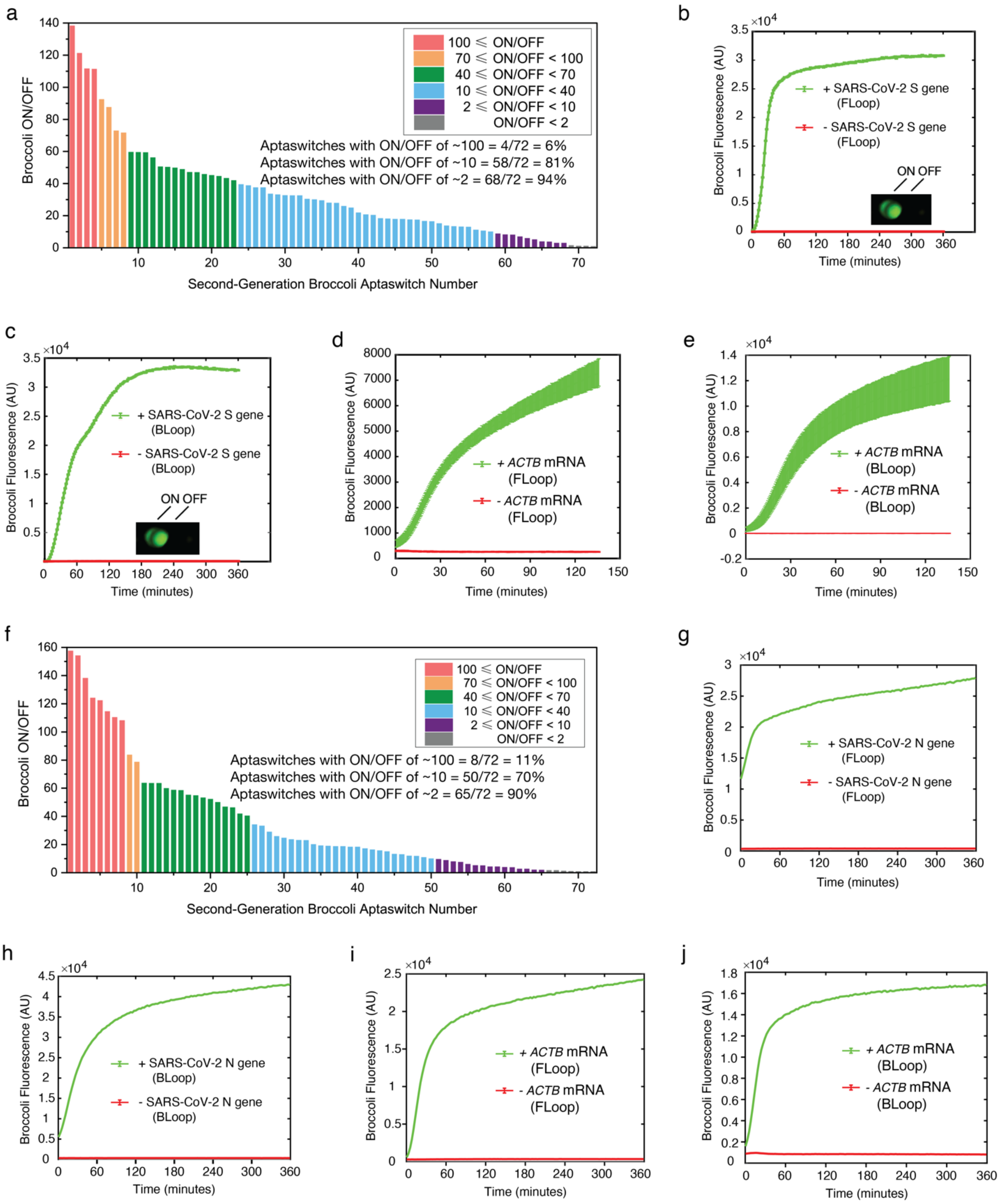
a
100 ⩽ ON/OFF
70 ⩽ ON/OFF < 100
40 ⩽ ON/OFF < 70
10 ⩽ ON/OFF < 40
2 ⩽ ON/OFF < 10
ON/OFF < 2
Aptaswitches with ON/OFF of ~100 = 4/72 = 6%
Aptaswitches with ON/OFF of ~10 = 58/72 = 81%
Aptaswitches with ON/OFF of ~2 = 68/72 = 94%
Broccoli ON/OFF
Second-Generation Broccoli Aptaswitch Number
b
+ SARS-CoV-2 S gene (FLoop)
- SARS-CoV-2 S gene (FLoop)
ON OFF
Broccoli Fluorescence (AU)
Time (minutes)
c
+ SARS-CoV-2 S gene (BLoop)
- SARS-CoV-2 S gene (BLoop)
ON OFF
Broccoli Fluorescence (AU)
Time (minutes)
d
+ ACTB mRNA (FLoop)
- ACTB mRNA (FLoop)
Broccoli Fluorescence (AU)
Time (minutes)
e
+ ACTB mRNA (BLoop)
- ACTB mRNA (BLoop)
Broccoli Fluorescence (AU)
Time (minutes)
f
100 ⩽ ON/OFF
70 ⩽ ON/OFF < 100
40 ⩽ ON/OFF < 70
10 ⩽ ON/OFF < 40
2 ⩽ ON/OFF < 10
ON/OFF < 2
Aptaswitches with ON/OFF of ~100 = 8/72 = 11%
Aptaswitches with ON/OFF of ~10 = 50/72 = 70%
Aptaswitches with ON/OFF of ~2 = 65/72 = 90%
Broccoli ON/OFF
Second-Generation Broccoli Aptaswitch Number
g
+ SARS-CoV-2 N gene (FLoop)
- SARS-CoV-2 N gene (FLoop)
Broccoli Fluorescence (AU)
Time (minutes)
h
+ SARS-CoV-2 N gene (BLoop)
- SARS-CoV-2 N gene (BLoop)
Broccoli Fluorescence (AU)
Time (minutes)
i
+ ACTB mRNA (FLoop)
- ACTB mRNA (FLoop)
Broccoli Fluorescence (AU)
Time (minutes)
j
+ ACTB mRNA (BLoop)
- ACTB mRNA (BLoop)
Broccoli Fluorescence (AU)
Time (minutes)

**Supplementary Fig. 5 I *In vitro* characterization of second-generation Broccoli aptaswitches for SARS-CoV-2 detection.**

**a**, ON/OFF fluorescence levels obtained for 72 Broccoli aptaswitches with the DFHBI-1T fluorogen determined in the presence or absence of the cognate stem-loop DNA that mimics the secondary structure of RT-LAMP DNA amplicons after a 2-hr reaction. These aptaswitches are designed for SARS-CoV-2 RNA or human control RNA (*ACTB* mRNA or 18s rRNA). Relative errors for the switch ON/OFF ratios were obtained by adding the relative errors of the switch ON and OFF state fluorescence measurements in quadrature. Relative errors for ON and OFF states are from the SD of *n*=3 technical replicates.

**b**-**e**, Time-course measurements of fluorescence with DFHBI-1T from four high-performance Broccoli aptaswitches for the detection of forward loop (**b**) and back loop (**c**) of SARS-CoV-2 RT-LAMP amplicon and forward loop (**d**) as well as back loop (**e**) of human control *ACTB* mRNA RT-LAMP amplicon.

**f**, ON/OFF fluorescence levels obtained for same Broccoli aptaswitches with the BI fluorogen determined in the presence or absence of the cognate stem-loop DNA that mimics the secondary structure of RT-LAMP DNA amplicons after a 2-hr reaction. Relative errors for the switch ON/OFF ratios were obtained by adding the relative errors of the switch ON and OFF state fluorescence measurements in quadrature.

**g**-**j**, Time-course measurements of fluorescence with BI fluorogen from four high-performance Broccoli aptaswitches for the detection of forward loop (**g**) and back loop (**h**) of SARS-CoV-2 RT-LAMP amplicon and forward loop (**i**) as well as back loop (**j**) of human control *ACTB* mRNA RT-LAMP amplicon.

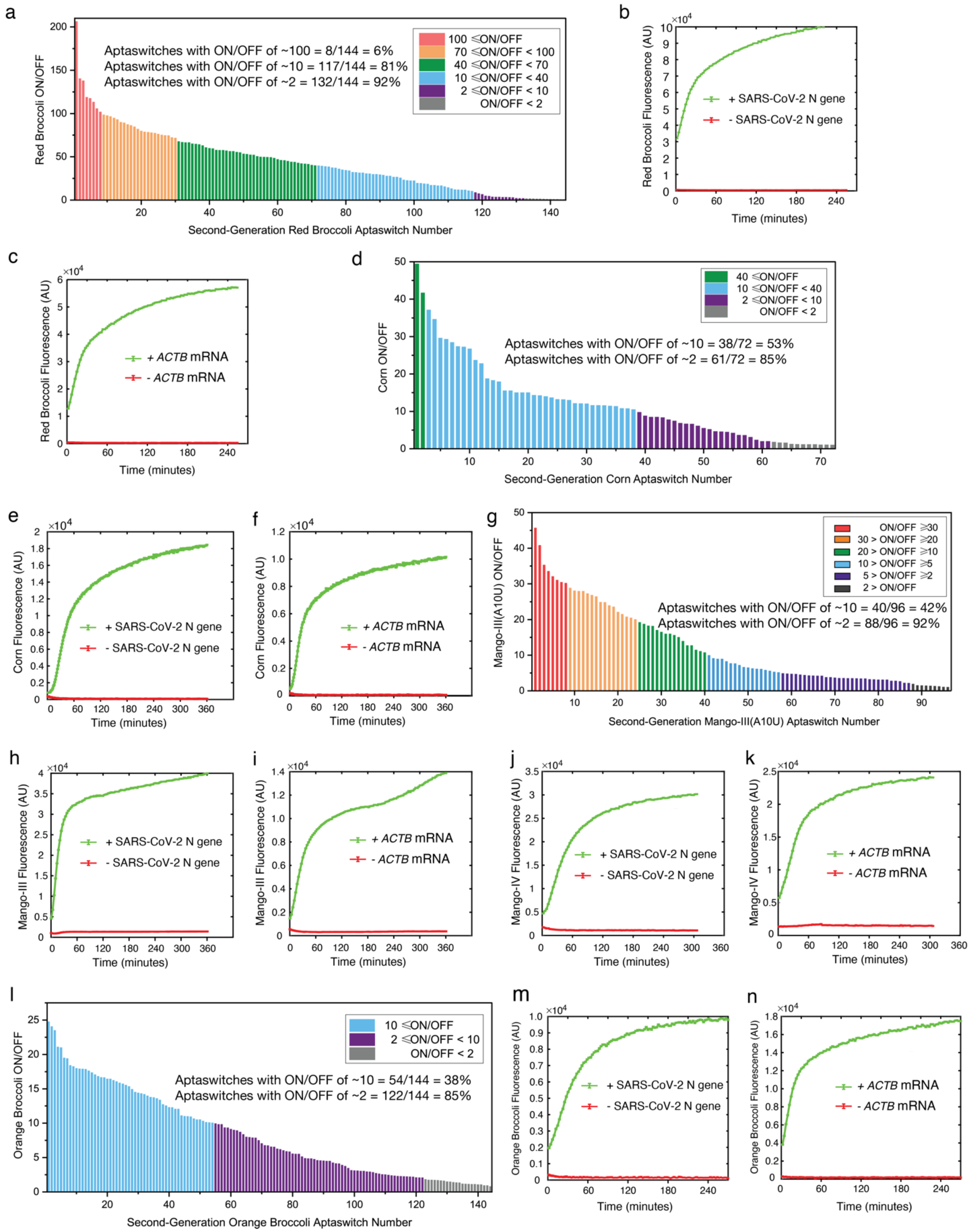


**Supplementary Fig. 6 | *In vitro* characterization of second-generation aptaswitches with**

**different aptamer outputs for SARS-CoV-2 RNA detection.**

**a**, **d**, **g**, and **l**, ON/OFF fluorescence levels obtained for 144 Red Broccoli aptaswitches (**a**), 72 Corn aptaswitches (**d**), 96 Mango-III(A10U) aptaswitches (**g**), and 144 Orange Broccoli aptaswitches (**l**) determined in the presence or absence of the cognate stem-loop DNA that mimics the secondary structure of RT-LAMP DNA amplicons after a 2-hr reaction. These aptaswitches are designed for SARS-CoV-2 RNA or human control RNA (*ACTB* mRNA or 18s rRNA). Relative errors for the switch ON/OFF ratios were obtained by adding the relative errors of the switch ON and OFF state fluorescence measurements in quadrature. Relative errors for ON and OFF states are from the SD of $n$=3 technical replicates.

**b**, **c**, **e**, **f**, **h**, **i**, **j**, **k**, **m**, and **n**, Time-course measurements of fluorescence from four high-performance aptaswitches for the detection SARS-CoV-2 RNA and human control *ACTB* mRNA using Red Broccoli (**b** and **c**), Corn (**e** and **f**), Mango-III(A10U) (**h** and **i**), Mango-IV (**j** and **k**), and Orange Broccoli (**m** and **n**) aptamer outputs, respectively.

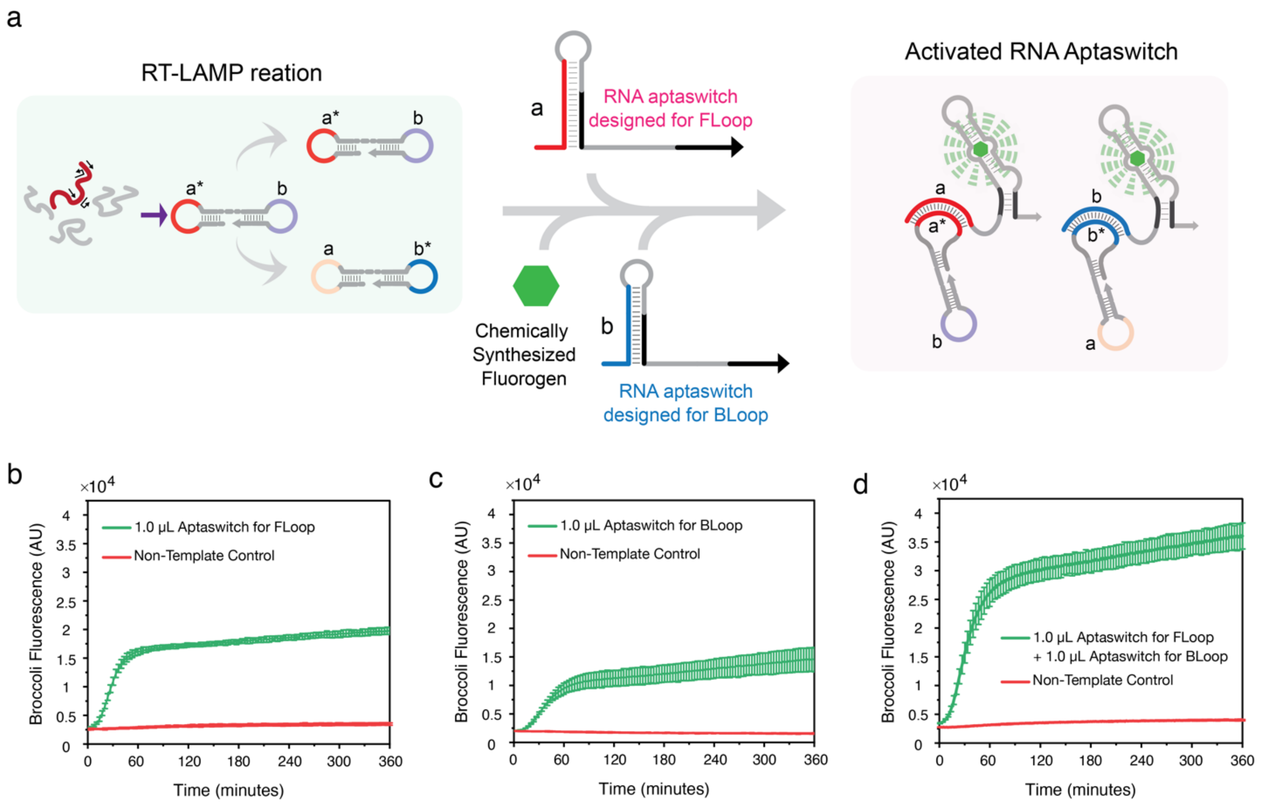


**Supplementary Fig. 7 I A dual-site targeting strategy for promoting the aptaswitch signal from the detection of RT-LAMP DNA amplicons.**

**a**, Scheme for parallel detection of two different RT-LAMP DNA loop domains in the same reaction. One aptaswitch targets the **a*** loop domain while the second aptaswitch targets the **b*** loop domain, enabling increases in signal and/or reaction speed.

**b-d**, Fluorescence measurements from validation of the dual loop detection scheme. On their own, aptaswitches for the FLoop (**b**) and BLoop (**c**) of the RT-LAMP DNA amplicon produce a fluorescence signal of 16,000 to 10,000 within a one-hour reaction period, respectively. In comparison, the dual loop system using aptaswitches targeting both the FLoop and BLoop simultaneously (**d**) provides roughly the sum of the two independent signals at ~26,000 after one hour. The increase in signal leads to a concomitant decrease in time-to-result of ~35%.

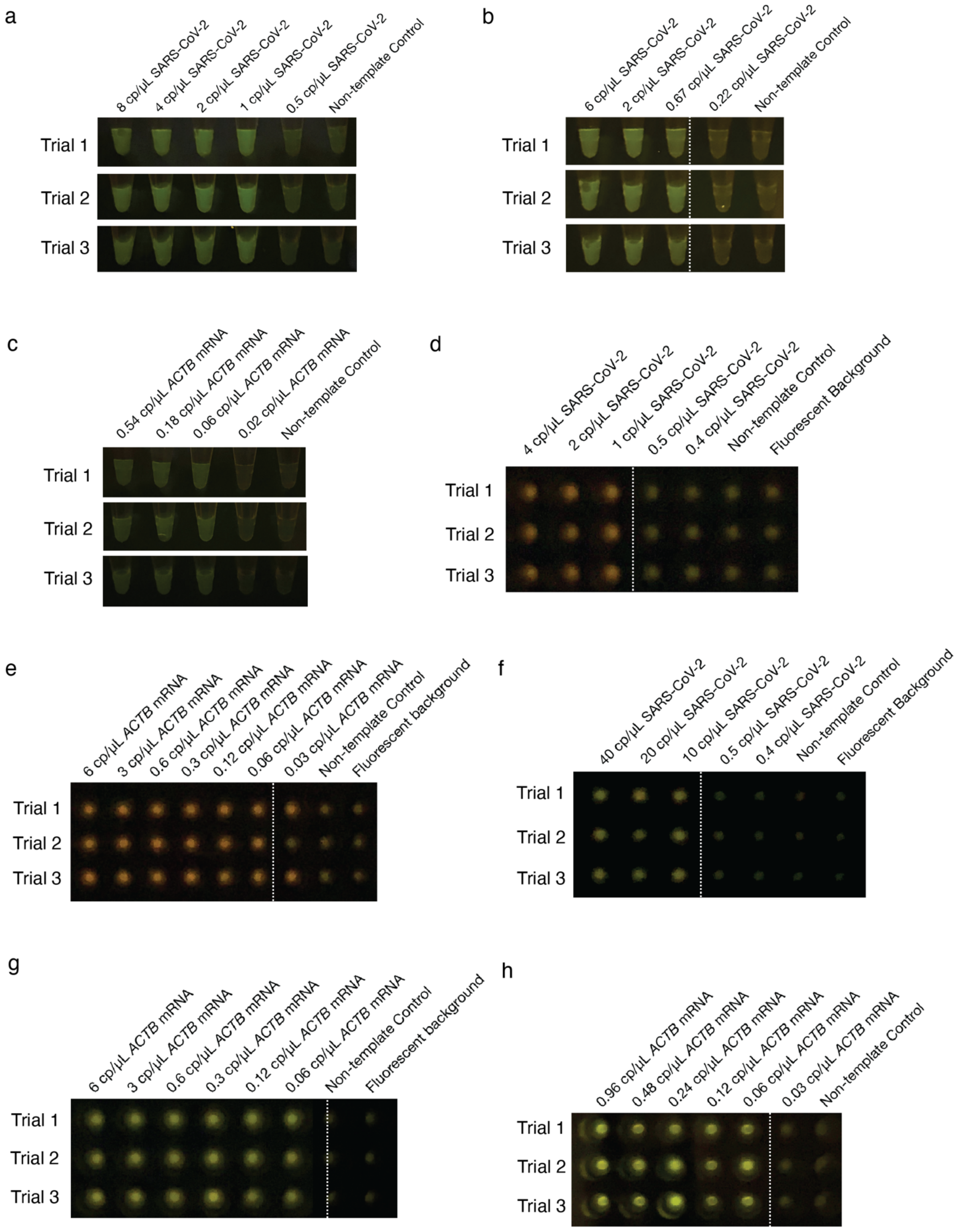
a
8 cp/μL SARS-CoV-2
4 cp/μL SARS-CoV-2
2 cp/μL SARS-CoV-2
1 cp/μL SARS-CoV-2
0.5 cp/μL SARS-CoV-2
Non-template Control
Trial 1
Trial 2
Trial 3
b
6 cp/μL SARS-CoV-2
2 cp/μL SARS-CoV-2
0.67 cp/μL SARS-CoV-2
0.22 cp/μL SARS-CoV-2
Non-template Control
Trial 1
Trial 2
Trial 3
c
0.54 cp/μL ACTB mRNA
0.18 cp/μL ACTB mRNA
0.06 cp/μL ACTB mRNA
0.02 cp/μL ACTB mRNA
Non-template Control
Trial 1
Trial 2
Trial 3
d
4 cp/μL SARS-CoV-2
2 cp/μL SARS-CoV-2
1 cp/μL SARS-CoV-2
0.5 cp/μL SARS-CoV-2
0.4 cp/μL SARS-CoV-2
Non-template Control
Fluorescent Background
Trial 1
Trial 2
Trial 3
e
6 cp/μL ACTB mRNA
3 cp/μL ACTB mRNA
0.6 cp/μL ACTB mRNA
0.3 cp/μL ACTB mRNA
0.12 cp/μL ACTB mRNA
0.06 cp/μL ACTB mRNA
0.03 cp/μL ACTB mRNA
Non-template Control
Fluorescent background
Trial 1
Trial 2
Trial 3
f
40 cp/μL SARS-CoV-2
20 cp/μL SARS-CoV-2
10 cp/μL SARS-CoV-2
0.5 cp/μL SARS-CoV-2
0.4 cp/μL SARS-CoV-2
Non-template Control
Fluorescent Background
Trial 1
Trial 2
Trial 3
g
6 cp/μL ACTB mRNA
3 cp/μL ACTB mRNA
0.6 cp/μL ACTB mRNA
0.3 cp/μL ACTB mRNA
0.12 cp/μL ACTB mRNA
0.06 cp/μL ACTB mRNA
Non-template Control
Fluorescent background
Trial 1
Trial 2
Trial 3
h
0.96 cp/μL ACTB mRNA
0.48 cp/μL ACTB mRNA
0.24 cp/μL ACTB mRNA
0.12 cp/μL ACTB mRNA
0.06 cp/μL ACTB mRNA
0.03 cp/μL ACTB mRNA
Non-template Control
Trial 1
Trial 2
Trial 3

**Supplementary Fig. 8 | Photograph of fluorescence from aptaswitch reactions following RT-LAMP of different SARS-CoV-2 RNA or human control *ACTB* mRNA concentrations.**

**a**-**c**, Strong green fluorescence observed from Broccoli aptaswitches designed for SARS-CoV-2 N gene (**a**), S gene (**b**) and human control *ACTB* mRNA (**c**). Reactions were measured in triplicate.

**d**-**e**, Strong red fluorescence observed from Red Broccoli aptaswitches designed for SARS-CoV-2 N gene (**d**) and human control *ACTB* mRNA (**e**). Reactions were measured in triplicate.

**f**-**g**, Strong orange fluorescence observed from Orange Broccoli aptaswitches designed for SARS-CoV-2 N gene (**f**) and human control *ACTB* mRNA (**g**). Reactions were measured in triplicate.

**h**, Strong yellow fluorescence observed from Mango-III(A10U) aptaswitches designed for human control *ACTB* mRNA. Reactions were measured in triplicate.

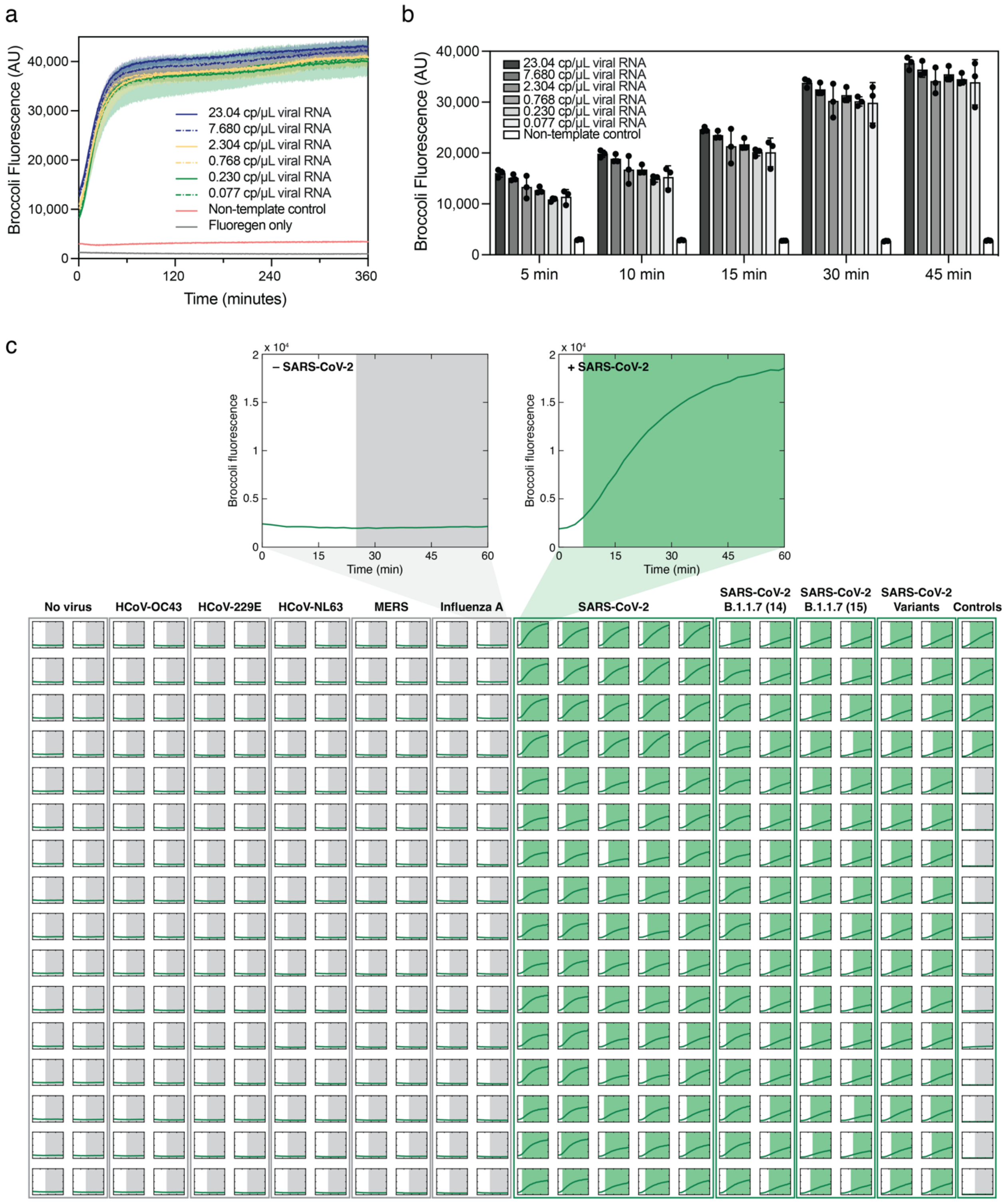


**Supplementary Fig. 9 | High-throughput SARS-CoV-2 RNA screening assay in 384-well plates.**

**a**, Time-course measurements of fluorescence from an S gene Broccoli aptaswitch after amplification with RT-LAMP using different cultured wild-type SARS-CoV-2 RNA concentrations. Shaded regions denote the arithmetic mean ± SD for *n*=3 technical replicates.

**b**, Fluorescence signal from Broccoli aptaswitches targeting SARS-CoV-2 after amplification with RT-LAMP at 5 min, 10 min, 15 min, 30min and 45 min. (*n*=3 technical replicates; bars represent the arithmetic mean ± SD)

**c**, High-throughput SARS-CoV-2 assay in 384-well plates. The aptaswitch assay was prepared in 384-well plates using a stable master mix formulation. Contrived samples were prepared from extracted RNA using concentrations typical of clinical saliva samples. The upper part of the figure shows the time-course measurements of Broccoli fluorescence for a representative pair of positive and negative SARS-CoV-2 wells. The green shaded region indicates the time point at which the Broccoli fluorescence passed the fluorescence threshold for samples identified as positive. In contrast, the gray region indicates the time at which a sample is identified as negative based on its fluorescence being below the positive threshold level after 25 minutes. The lower part shows the Broccoli fluorescence readouts from all 384 wells on the plate. Control samples occupy the far-right column, while negative samples occupy the left half of the plate. The remaining wells, occupying most of the right side of the plate, contain RNA from different SARS-CoV-2 strains. Rapid response of the Broccoli aptaswitches against a variety of SARS-CoV-2 variants enabled positive calls to be made within 25 minutes of incubation in a plate reader. 176 out of 176 positive samples and 192 out of 192 negative samples were correctly identified. 16 wells (right-most column of plate) were used for controls. The assays did not activate in the presence of other human coronaviruses, MERS, and influenza A.

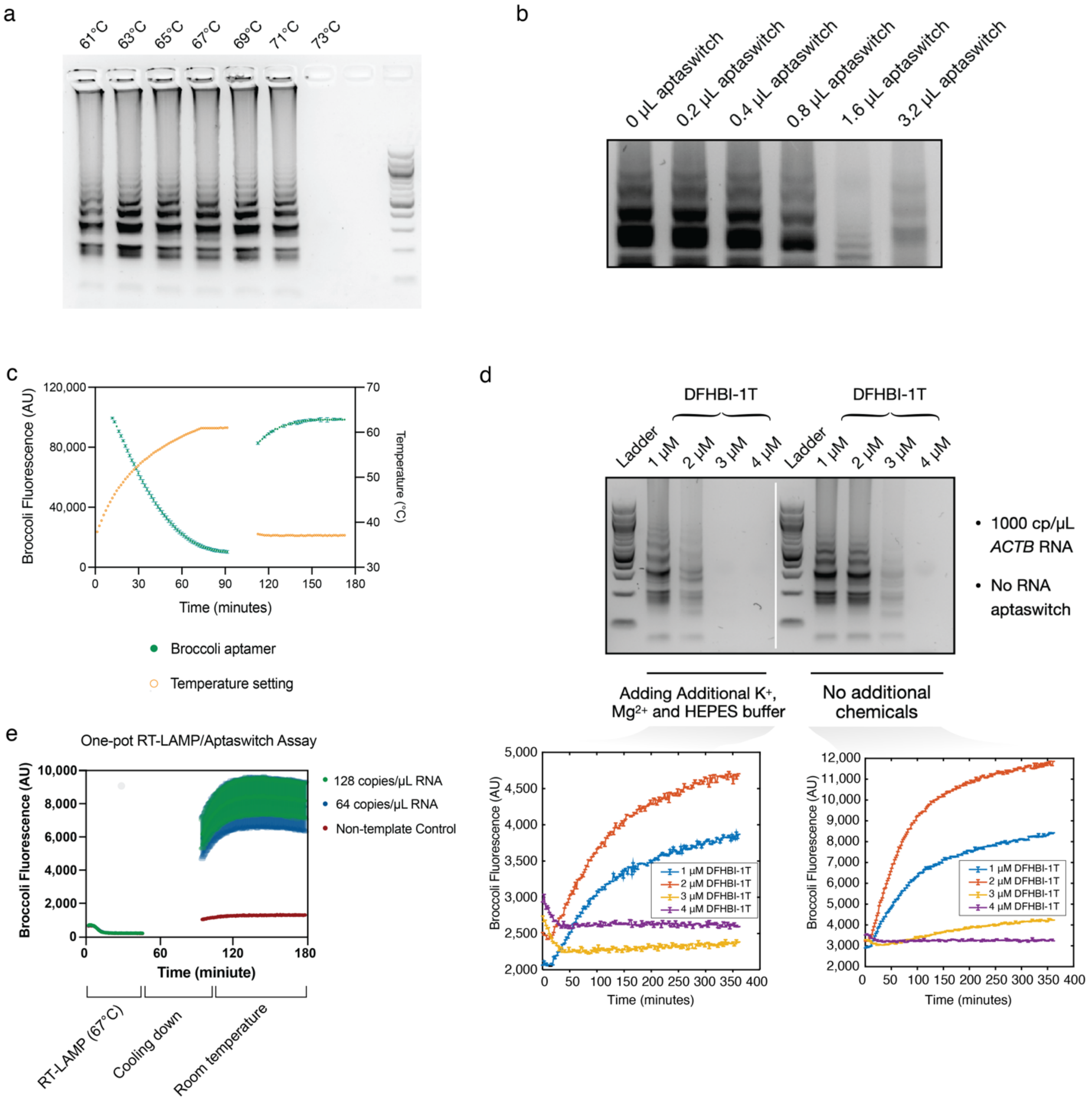


**Supplementary Fig. 10 I Temperature optimization for adapting aptaswitches to RT-LAMP for a one-pot diagnostic system.**

**a**, Functionality tests of RT-LAMP enzymes (Bst 2.0 WarmStart DNA Polymerase and WarmStart RTx Reverse Transcriptase) under different temperatures between 61°C and 73°C.

**b**, Functionality tests of RT-LAMP reactions when mixing with different volumes of transcribed RNA Broccoli Aptaswitch between 0.2 $\mu$L and 3.2 $\mu$L while keeping the concentration of RT-LAMP primers constant.

**c**, Fluorescence performance of Broccoli aptaswitches at 61°C and 37°C as a function of time and temperature. Much more fluorescence is observed at 37°C compared to 61°C.

**d**, Functionality tests of RT-LAMP enzymes (Bst 2.0 WarmStart DNA Polymerase and WarmStart RTx Reverse Transcriptase) under different concentrations of DFHBI-1T between 1 $\mu$M and 4 $\mu$M with (left) or without (right) 40 mM HEPES buffer, 100 mM KCl, and 5 mM $MgCl_2$. The amplification products were then diluted with Broccoli aptaswitches for fluorescence measurements (bottom).

**e**, Time-course measurements of fluorescence from dual S gene-targeted Broccoli aptaswitches in an RT-LAMP-coupled one-pot reaction. The reaction is first heated to 67°C for 45 minutes to amplify the SARS-CoV-2 RNA from the pathogen, then the temperature of the system is reduced enabling the proper folding of aptaswitches and aptaswitch binding to the amplicons generates a fluorescence signal indicating the presence of the target nucleic acids.

a

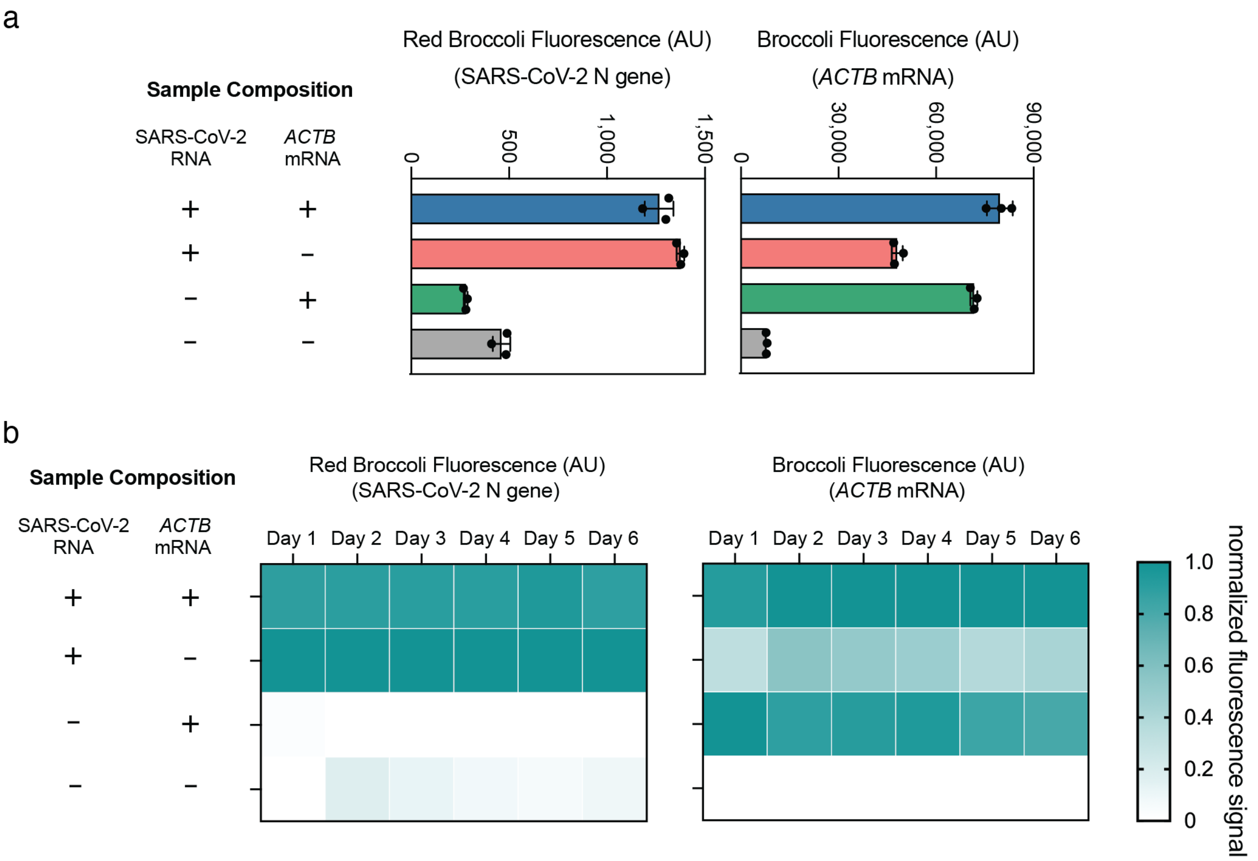


**Supplementary Fig. 11 I Stability tests of aptaswitch fluorescence signals.**

**b**, First-day fluorescence measurements of all-in-one one-pot two-channel detection of SARS-CoV-2 RNA and human control *ACTB* mRNA with Red Broccoli and Broccoli aptaswitches, respectively. (*n*=3 technical replicates; bars represent the arithmetic mean ± SD)

**c**, Measurements of aptaswitch fluorescence intensity over 6 days. The heat map represents the arithmetic mean of three replicates.

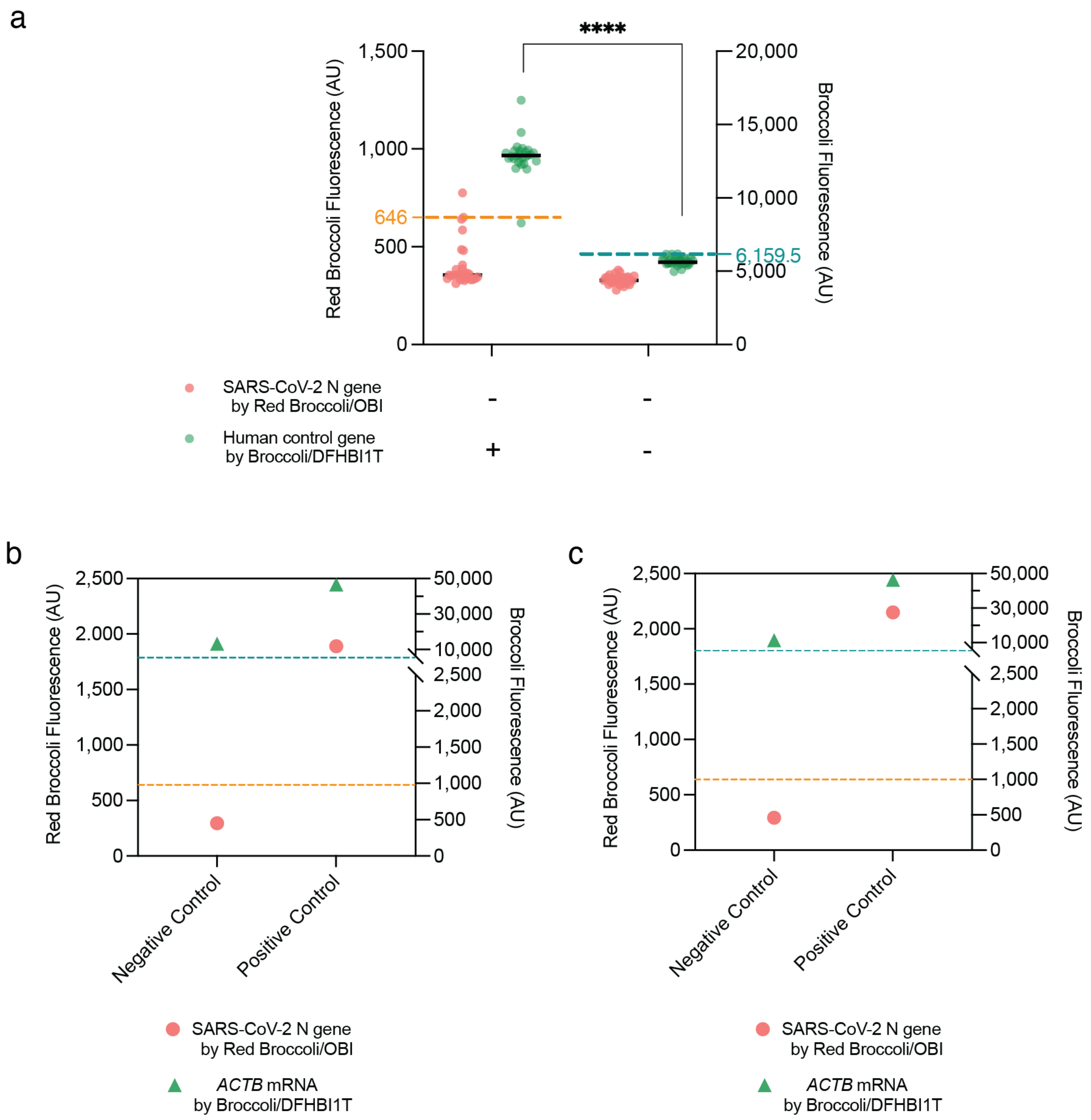


**Supplementary Fig. 12 | Control experiments for multiplexed one-pot RT-LAMP/aptaswitch assays with clinical samples.**

**a**, The 95$^{th}$ percentile fluorescence values after return to room temperature. Two sets of control samples were used: (1) one without SARS-CoV-2 RNA but with *ACTB* mRNA, and (2) one with neither SARS-CoV-2 RNA nor *ACTB* mRNA. 95$^{th}$ percentile fluorescence levels from these control samples were used define threshold levels for identification of SARS-CoV-2 RNA and *ACTB* mRNA in the samples.

**b**, Control experiment results for detection of extracted RNA from clinical saliva samples. The dashed line represents the diagnostic threshold required for sample assessment (orange dashed line: Red Broccoli aptaswitch threshold value for determining a positive sample; teal dashed line: Broccoli aptaswitch threshold value for determining a valid sample).

**c**, Control experiment results for one-pot extraction-free detection of clinical saliva samples. The dashed line represents the diagnostic threshold (orange dashed line: Red Broccoli aptaswitch threshold value for determining a positive sample; teal dashed line: Broccoli aptaswitch threshold value for determining a valid sample).
